# Lockdown babies- Birth and new parenting experiences during the 2020 Covid-19 lockdown in South Africa

**DOI:** 10.1101/2021.05.20.21257537

**Authors:** Elise Farley, Amanda Edwards, Emma Numanoglu, Tamsin K. Phillips

## Abstract

**Background:** Perceived birth experiences of parents can have a lasting impact on children. We explored the birth and new parenting experiences of South African parents during the Covid-19 lockdown.

**Methods:** We conducted an online cross-sectional survey with consenting parents of babies born in South Africa during 2020. Factors associated with negative birth emotions and probable depression were estimated using logistic regression.

**Results:** Most of the 520 respondents were females (n= 496, 95%) who gave birth at private hospitals (n=426, 86%). Mothers reported having overall positive birth emotions (n= 399, 80%). Multivariable analysis showed that having the baby during lockdown (adjusted odds ratio (aOR) 5.02; CI 1.28 – 19.66); being diagnosed with Covid-19 (aOR 3.17; CI 1.07 – 9.42); having negative new parenting emotions (aOR 6.07; CI 3.27 – 11.29); a preterm baby (aOR 3.02; CI 1.36 – 6.70) and lockdown related barring of preferred in hospital support (aOR 2.45; CI 1.35 – 4.43) were associated with mothers reporting predominately negative emotions about the birth. Having their chosen delivery method reduced the odds of negative birth emotions (aOR 0.4; CI 0.22 – 0.72). Multivariable analysis showed that having predominantly negative new parenting emotions (aOR 10.75; CI 5.41-21.37), breastfeeding struggles (aOR 2.16; CI 1.36 – 3.46); lockdown preventing health care access (aOR 2.06; CI 1.20 – 3.54) and creating financial strain (aOR 2.58; CI 1.08 – 6.18) were associated with probable minor depression

**Conclusions:** Lockdown exacerbated many birth and parenting challenges including mental health and health care access. However, overall experiences were positive and there was a strong sense of resilience amongst parents.

## Introduction

The first 1000 days of a baby’s life are the most crucial in terms of development [1]. As part of these 1000 days, the birth of a baby and the perceived birth experiences of the parents can have a lasting impact on the child’s life. Subjective birth-experiences have been shown to be associated with the development of post-traumatic stress symptoms in the mother [2, 3] and the mental health of parents can have a long-lasting impact on the child’s behavioural and emotional state [4, 5].

The Covid-19 pandemic and ensuing national lockdowns have had widespread global ramifications on every aspect of life, including population wide mental health [6, 7]. These “lockdowns” have severely altered people’s social networks and support structures due to the restrictions on seeing family and friends, drastic changes to routines and, for some, impacted economic livelihoods [7]. South Africa entered one of the world’s strictest lockdowns on 26 March 2020, and followed a tiered approach in attempts to curb the spread of COVID-19 nationally.

Both having a baby and living through lockdown are stressful situations where support is imperative to ensure positive outcomes for both parents and children. A United Kingdom based study assessing the maternal experiences and infant feeding during the Covid-19 period showed that lockdown has had an impact on parents, resulting in difficulties including changes in delivery plan, a lack of breastfeeding support, no maternal healthcare, mental health challenges and a lack of social support [8]. In a similar manner, we explored the unique birth and new parenting experiences in South Africa during the Covid-19 lockdown.

## Participants, Ethics and Methods

### Study design, setting and study participants

We conducted cross-sectional online survey with parents of babies born in South Africa during 2020. Data collection took place from 09 September to 10 October 2020. Participants were aged 18 years or older, living in South Africa at the time of the baby’s birth and had a baby born between 1 January 2020 until 10 October 2020.

### Sampling

A mixture of convenience and snowball sampling was used for the study. We initially sent the survey out to networks of parents who attended antenatal courses run by co-author EN (through her organisation Breastfeeding Matters). This grouping was chosen as it was anticipated that these parents had children born during our study period. The co-authors also sent the survey out on WhatsApp parenting support groups. Respondents were asked to share the survey with other parents who met the inclusion criteria. Due to the time sensitive nature of the study and restrictions in reaching potential respondents via alternative mechanisms under ongoing lockdown conditions, this recruitment strategy was chosen as it was thought to be the most appropriate for the respondent population who are frequently connected through multiple online formats.

### Data collection

The data collection tool was created in English using KoBoCollect (KoboToolBox©). A link to the survey was generated and sent out to respondents over email, WhatsApp and via social media. The questionnaire includes a selection of questions derived from a similar United Kingdom study [8] (Supplementary material, Table S1).

Data collected included sociodemographic characteristics, details of the birth and baby, information on feeding. Respondents were asked about their predominant emotions during birth and the new parenting phase. A selection of emotions were listed under the positive (joyful, grateful, relieved, hopeful) and negative (fearful, resentful, inadequate, anxious, sad, overwhelmed) response to guide respondents. Respondents were asked about how Covid-19 had impacted them. Both closed and open response questions were included to explore the full range of emotions experienced.

The 10 point Edinburgh Postnatal Depression Scale [9] was used to assess which respondents showed symptoms of probable depression. It must be noted that this tool was not used as a diagnostic tool. Minor depression was indicated for respondents who scored 10 or more and major depression was indicated for those who scored 13 or more [10]. Minor depression was used in our analysis as this cut off is more sensitive than that for major depression. Participants who scored above 10 were prompted to reach out for support and all participants were provided with resources for psychosocial support services.

### Analysis

Due to the small sample size of male respondents (n=24), we conducted separate analyses on responses from female and male respondents. For the male cohort, we performed a descriptive analyses. Categorical variables are reported as frequencies and proportions. Depending on their distribution, continuous variables are summarised using medians and interquartile ranges (IQR) or means and standard deviations (SD).

For the female cohort, we performed a similar descriptive analysis. In addition, we compared the responses between those who reported having predominately positive emotions about their birth, and those who had predominantly negative emotions about their birth. Comparisons were also made by lockdown level at the time of delivery (5 categories: Alert Level 5: 26 March - 30 April, Level 4: 1 – 31 May, Level 3: 1 June – 17 August, Level 2: 18 August – 20 September, Level 1: 21 September – 28 December, after which time adjusted levels were implemented), and in binary between those whose babies were born prior to lockdown (1 January – 25 March 2020) and those who babies were born during lockdown (25 March – end of data collection on 10 October 2020). Bivariate associations were examines using chi-squared tests, t-tests and Kruskal Wallis tests as appropriate. Missing data numbers are recorded in each table.

Univariable and multivariable analysis were conducted using logistic regression, to estimate factors associated with predominant birth emotions (positive and negative) and probable minor depression (yes or no). Variables chosen for inclusion in the multivariable analysis were those with ten or more cases and a univariable strength of association equivalent to a p-value < 0.2, after assessing collinearity among variables [11]. Odds ratios (OR), adjusted odds ratios (aOR) and their respective 95% confidence intervals (CI) are reported. All quantitative data analysis was conducted with Stata 15 (StataCorp LP, College Station, TX, USA).

To complement quantitative analyses, free text responses were analysed by coinvestigator AE using thematic content analysis [5]. Coding was conducted by hand, allowing for the addition of new concepts in two rounds of analysis. The frequency of each concept was then catalogued and quantified in MS Excel, before high-frequency related concepts were merged into core themes.

### Ethical statement

Ethical approval was obtained from the University of Cape Town Human Research Ethics Committee (385/2020). Informed consent was obtained from all respondents by asking them to first read the information sheet and then provide consent by selecting the ‘Yes’ button on the online survey. If respondents declined to participate after reading the information sheet, this was recorded and they were directed to a page with parenting and mental health resources. The survey was anonymous and no identifying information such as names or addresses were collected.

## Results

In total, 520 parents completed the survey, 496 (95%) were females and 24 (5%) were males.

### Females

#### Socio-Demographic characteristics

The mean age of female respondents was 32 years (SD 4 years); 327 (66%) were from the Western Cape; 183 (37%) highest qualification was an undergraduate degree; 339 (68% were employed full-time at the time of the survey; 474 (96%) were married, in a civil partnership or cohabiting with a partner and 426 (86%) gave birth at a private hospital (Table 1). Mothers in our study gave birth to 506 babies in total, 255 were males (50%). For just over half (n=260; 53%) of the mothers in the study, this was their first child. The majority were born at a normal gestational age range (37-42 completed weeks of pregnancy [12], n=436, 88%). The mean birth weight was 3.2kg (SD 0.6kg), 47 (10%) babies were classified as having a low birth weight (0-2499g), 414 (84%) as normal birthweight (2500 – 3999g) and 32 (7%) as having a high birthweight (>4000g).

**Table 1.**
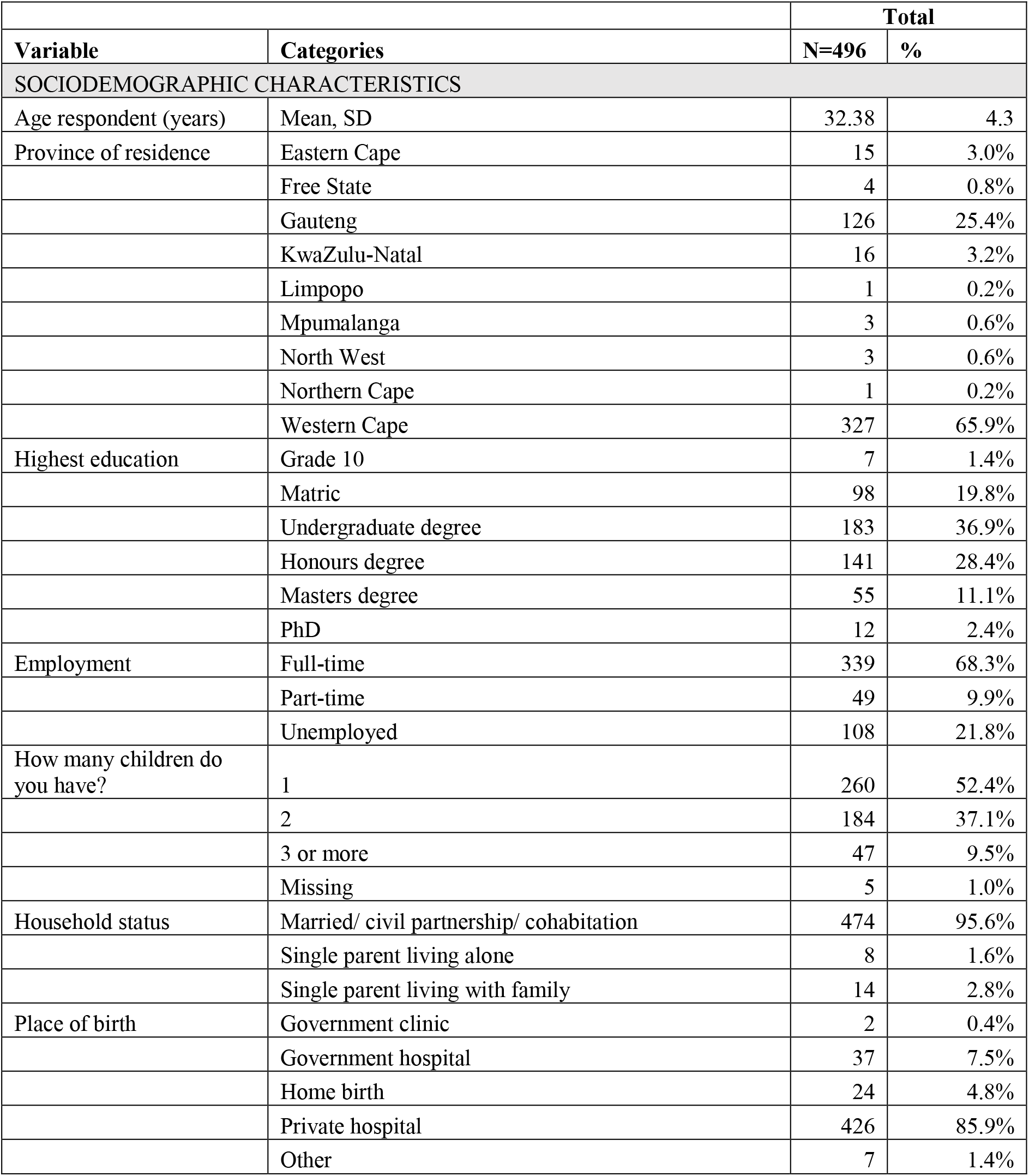
Female respondent’s socio-demographic characteristics and the impact of Covid-19 lockdown.

|  |  | **Total** |  |
| --- | --- | --- | --- |
| **Variable** | **Categories** | **N=496** | **%** |
| **SOCIODEMOGRAPHIC CHARACTERISTICS** |  |  |  |
| Age respondent (years) | Mean, SD | 32.38 | 4.3 |
| Province of residence | Eastern Cape | 15 | 3.0% |
|  | Free State | 4 | 0.8% |
|  | Gauteng | 126 | 25.4% |
|  | KwaZulu-Natal | 16 | 3.2% |
|  | Limpopo | 1 | 0.2% |
|  | Mpumalanga | 3 | 0.6% |
|  | North West | 3 | 0.6% |
|  | Northern Cape | 1 | 0.2% |
|  | Western Cape | 327 | 65.9% |
| Highest education | Grade 10 | 7 | 1.4% |
|  | Matric | 98 | 19.8% |
|  | Undergraduate degree | 183 | 36.9% |
|  | Honours degree | 141 | 28.4% |
|  | Masters degree | 55 | 11.1% |
|  | PhD | 12 | 2.4% |
| Employment | Full-time | 339 | 68.3% |
|  | Part-time | 49 | 9.9% |
|  | Unemployed | 108 | 21.8% |
| How many children do you have? | 1 | 260 | 52.4% |
|  | 2 | 184 | 37.1% |
|  | 3 or more | 47 | 9.5% |
|  | Missing | 5 | 1.0% |
| Household status | Married/ civil partnership/ cohabitation | 474 | 95.6% |
|  | Single parent living alone | 8 | 1.6% |
|  | Single parent living with family | 14 | 2.8% |
| Place of birth | Government clinic | 2 | 0.4% |
|  | Government hospital | 37 | 7.5% |
|  | Home birth | 24 | 4.8% |
|  | Private hospital | 426 | 85.9% |
|  | Other | 7 | 1.4% |

#### Lockdown impact

The Covid-19 lockdown led to mothers reporting delays in registering their babies (n=231, 47%) and difficulties in purchasing baby goods (n=229, 46%). The main non-medical activities that mothers missed due to lockdown were visiting friends and family (n=441, 89%), outdoor exercise (n=197, 40%) and going to restaurants (n=179, 36%). The majority of babies in the study cohort were born during lockdown (pre-lockdown n=78, 16%; level 5 n=95, 19%; level 4 n=88, 18%; level 3 n= 168, 34%; level 2 n= 64, 13% and level 1 n=3, 1%). Mothers with babies born prior to lockdown were more likely to have their preferred method of birth (n=51; 65%) in comparison to those with babies born during level 4 (n=48; 55), level 3 (n=97; 58%) and level 2 (n=37; 58%) lockdown (p=0.009). There were fewer babies placed skin to skin after birth during lockdown (pre-lockdown n=67, 86%; lockdown n= 313, 75%; p=0.040). More mothers reported having a negative birth experience during lockdown (pre-lockdown n=3, 4%; lockdown n= 94, 23%; p=<0.001). A vast majority of mothers who gave birth during lockdown reported that the pandemic had affected their birth experience (pre-lockdown n=16, 21%; lockdown n= 325, 78%; p=<0.001) and nearly half reported that someone they had wanted to be at the birth was not able to be there because of the Covid-19 pandemic (pre-lockdown n=6, 8%; lockdown n= 198, 47%; p=<0.001). A third as many mothers saw a lactation consultant in hospital during lockdown (n=53, 13%) in comparison to prior to lockdown (n=30, 39%; p=<0.001). Lockdown did not affect the number of babies born preterm (pre-lockdown n=9, 12%; lockdown n=48, 12%) (Supplementary material, Table S2).

#### Regression

##### Predominant emotions about birth

The majority of mothers reported having overall positive emotions about their birth (positive n= 399, 80%; negative n=97, 20%). The main reasons for negative birth experiences were a traumatic experience (n=42, 8%), feeling disconnected from what was happening (n= 37, 7%), and not having a partner at the birth (n=34, 7%). Figure 1 illustrates the words mothers used most commonly to describe their birth experience. The risk factors associated with having predominately negative emotions at birth are shown in Table 2. These include having the baby during lockdown (aOR 5.02; CI 1.28 – 19.66); being diagnosed with Covid-19 (aOR 3.17; CI 1.07 – 9.42); having predominantly negative emotions during the new parenting phase (aOR 6.07; CI 3.27 – 11.29); having a preterm baby (aOR 3.02; CI 1.36 – 6.70) and lockdown related barring of preferred in hospital support for the birth (aOR 2.45; CI 1.35 – 4.43). Being able to have their delivery method reduced the odds of having negative birth emotions (aOR 0.4; CI 0.22 – 0.72) (Table 2).

**Figure 1:**
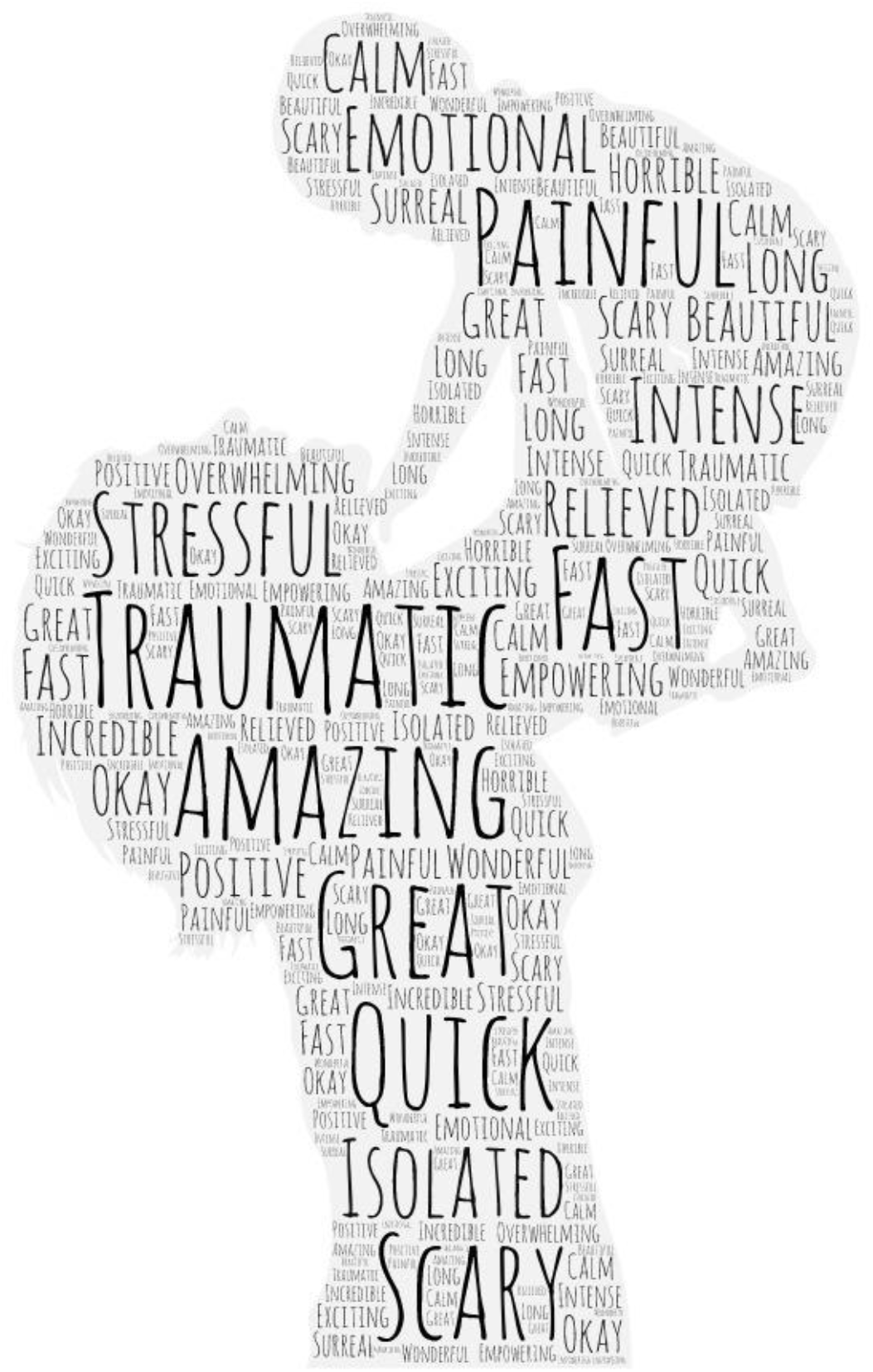
Words mothers used to describe their birth experience

**Table 2:**
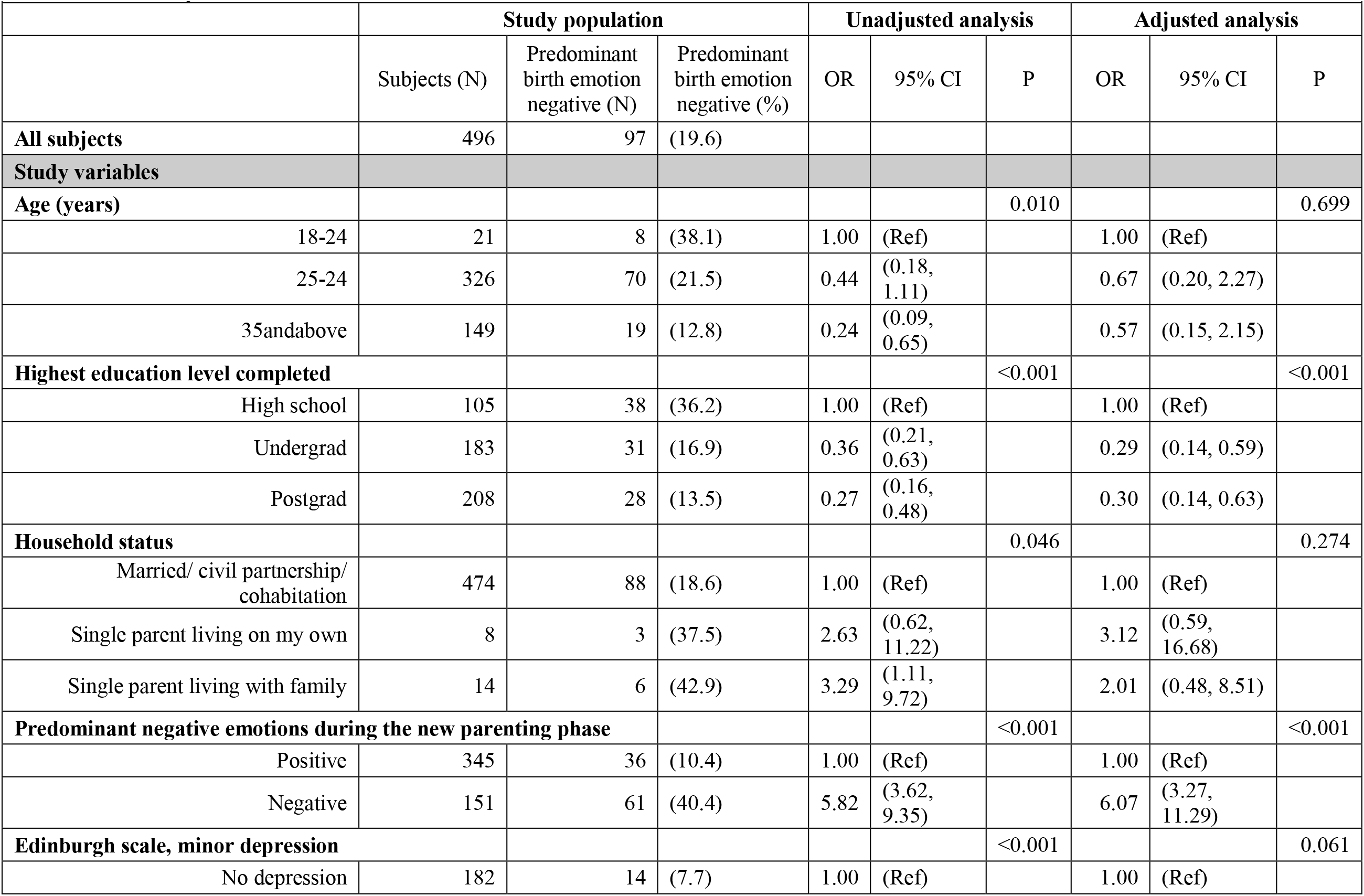
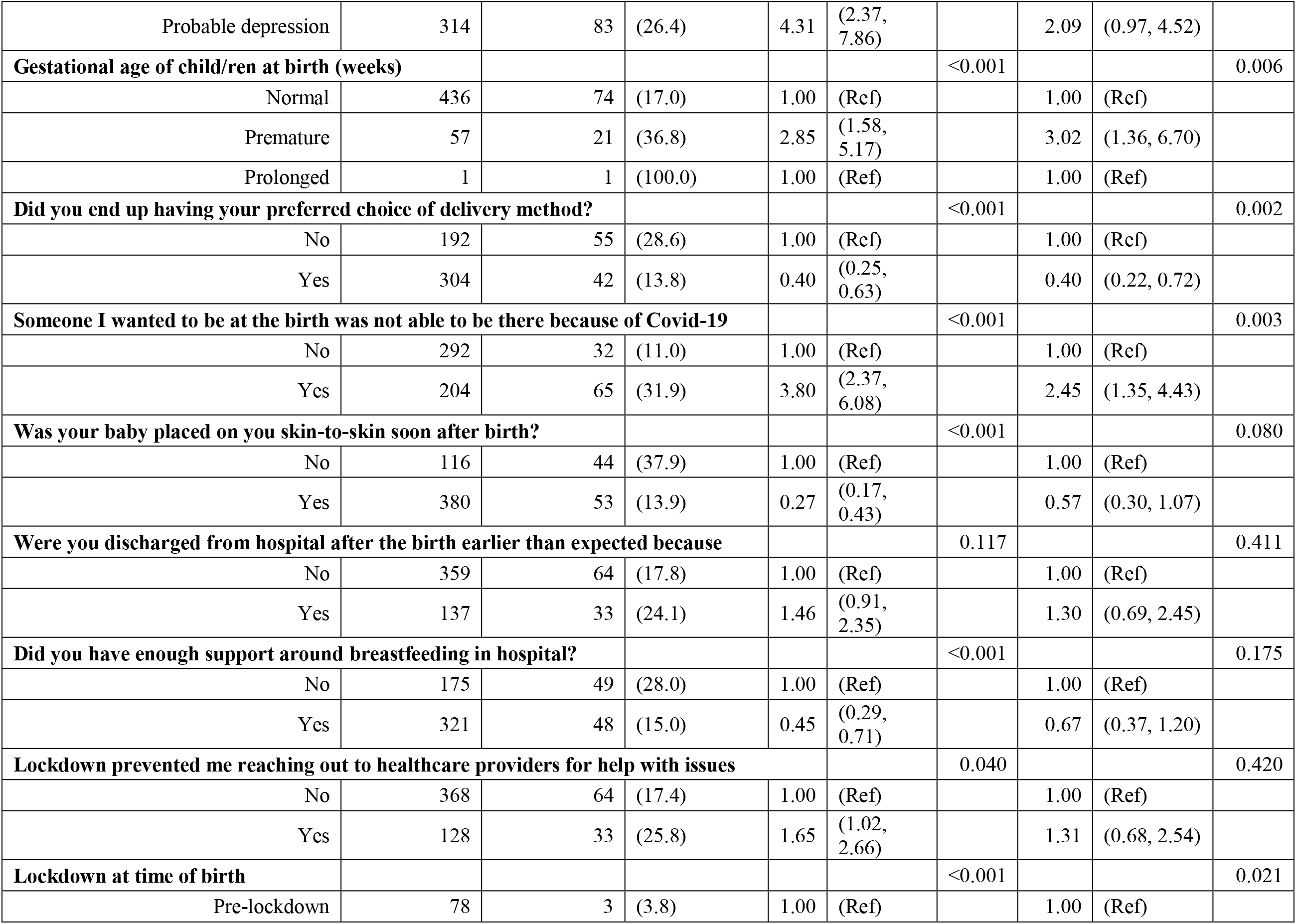
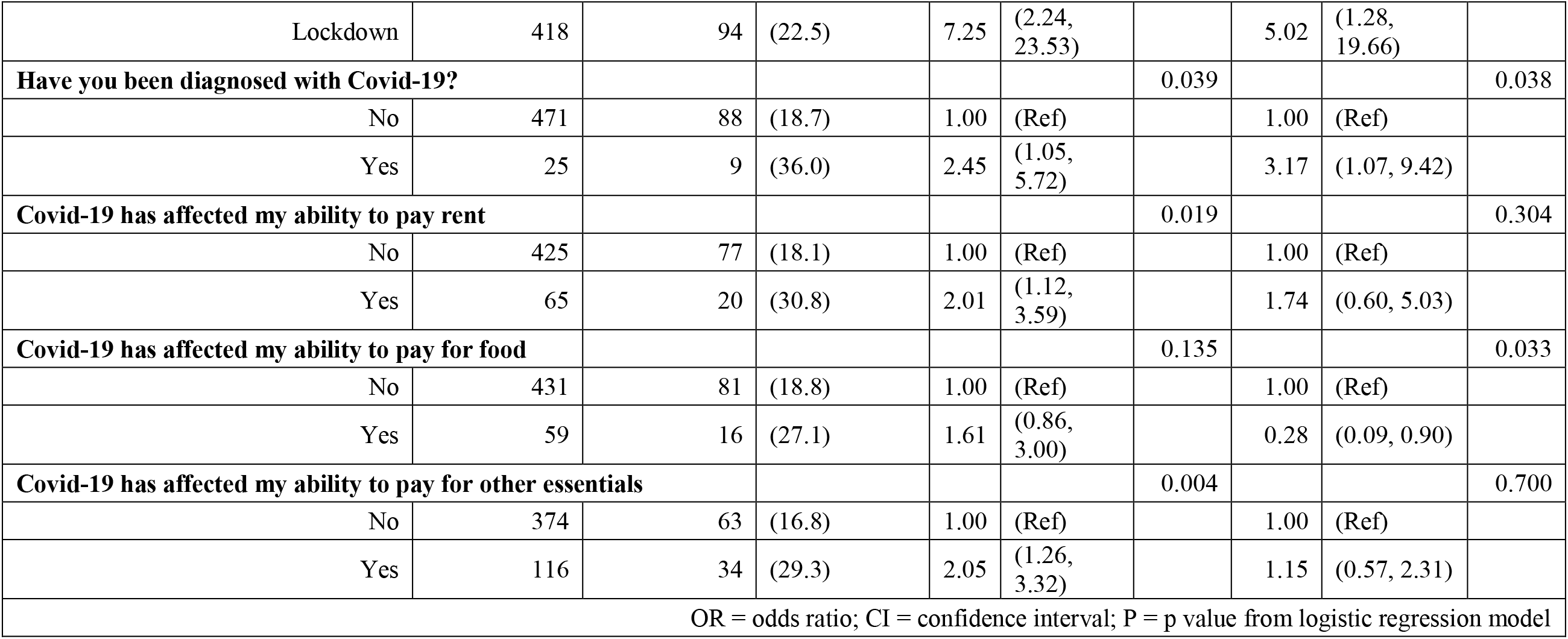
Factors associated with predominantly negative emotions about the birth, variables with p<0.2 in the univariable analysis include in multivariable analysis.

|  |  |  |  |  |  |  |  |  |  |
| --- | --- | --- | --- | --- | --- | --- | --- | --- | --- |
|  | Study population |  |  | Unadjusted analysis |  |  | Adjusted analysis |  |  |
|  | Subjects (N) | Predominant birth emotion negative (N) | Predominant birth emotion negative (%) | OR | 95% CI | P | OR | 95% CI | P |
| **All subjects** | 496 | 97 | (19.6) |  |  |  |  |  |  |
| **Study variables** |  |  |  |  |  |  |  |  |  |
| **Age (years)** |  |  |  |  |  | 0.010 |  |  | 0.699 |
| 18-24 | 21 | 8 | (38.1) | 1.00 | (Ref) |  | 1.00 | (Ref) |  |
| 25-24 | 326 | 70 | (21.5) | 0.44 | (0.18, 1.11) |  | 0.67 | (0.20, 2.27) |  |
| 35andabove | 149 | 19 | (12.8) | 0.24 | (0.09, 0.65) |  | 0.57 | (0.15, 2.15) |  |
| **Highest education level completed** |  |  |  |  |  | <0.001 |  |  | <0.001 |
| High school | 105 | 38 | (36.2) | 1.00 | (Ref) |  | 1.00 | (Ref) |  |
| Undergrad | 183 | 31 | (16.9) | 0.36 | (0.21, 0.63) |  | 0.29 | (0.14, 0.59) |  |
| Postgrad | 208 | 28 | (13.5) | 0.27 | (0.16, 0.48) |  | 0.30 | (0.14, 0.63) |  |
| **Household status** |  |  |  |  |  | 0.046 |  |  | 0.274 |
| Married/ civil partnership/ cohabitation | 474 | 88 | (18.6) | 1.00 | (Ref) |  | 1.00 | (Ref) |  |
| Single parent living on my own | 8 | 3 | (37.5) | 2.63 | (0.62, 11.22) |  | 3.12 | (0.59, 16.68) |  |
| Single parent living with family | 14 | 6 | (42.9) | 3.29 | (1.11, 9.72) |  | 2.01 | (0.48, 8.51) |  |
| **Predominant negative emotions during the new parenting phase** |  |  |  |  |  | <0.001 |  |  | <0.001 |
| Positive | 345 | 36 | (10.4) | 1.00 | (Ref) |  | 1.00 | (Ref) |  |
| Negative | 151 | 61 | (40.4) | 5.82 | (3.62, 9.35) |  | 6.07 | (3.27, 11.29) |  |
| **Edinburgh scale, minor depression** |  |  |  |  |  | <0.001 |  |  | 0.061 |
| No depression | 182 | 14 | (7.7) | 1.00 | (Ref) |  | 1.00 | (Ref) |  |
| Probable depression | 314 | 83 | (26.4) | 4.31 | (2.37, 7.86) |  | 2.09 | (0.97, 4.52) |  |
| **Gestational age of child/ren at birth (weeks)** |  |  |  |  |  | <0.001 |  |  | 0.006 |
| Normal | 436 | 74 | (17.0) | 1.00 | (Ref) |  | 1.00 | (Ref) |  |
| Premature | 57 | 21 | (36.8) | 2.85 | (1.58, 5.17) |  | 3.02 | (1.36, 6.70) |  |
| Prolonged | 1 | 1 | (100.0) | 1.00 | (Ref) |  | 1.00 | (Ref) |  |
| **Did you end up having your preferred choice of delivery method?** |  |  |  |  |  | <0.001 |  |  | 0.002 |
| No | 192 | 55 | (28.6) | 1.00 | (Ref) |  | 1.00 | (Ref) |  |
| Yes | 304 | 42 | (13.8) | 0.40 | (0.25, 0.63) |  | 0.40 | (0.22, 0.72) |  |
| **Someone I wanted to be at the birth was not able to be there because of Covid-19** |  |  |  |  |  | <0.001 |  |  | 0.003 |
| No | 292 | 32 | (11.0) | 1.00 | (Ref) |  | 1.00 | (Ref) |  |
| Yes | 204 | 65 | (31.9) | 3.80 | (2.37, 6.08) |  | 2.45 | (1.35, 4.43) |  |
| **Was your baby placed on you skin-to-skin soon after birth?** |  |  |  |  |  | <0.001 |  |  | 0.080 |
| No | 116 | 44 | (37.9) | 1.00 | (Ref) |  | 1.00 | (Ref) |  |
| Yes | 380 | 53 | (13.9) | 0.27 | (0.17, 0.43) |  | 0.57 | (0.30, 1.07) |  |
| **Were you discharged from hospital after the birth earlier than expected because** |  |  |  |  |  | 0.117 |  |  | 0.411 |
| No | 359 | 64 | (17.8) | 1.00 | (Ref) |  | 1.00 | (Ref) |  |
| Yes | 137 | 33 | (24.1) | 1.46 | (0.91, 2.35) |  | 1.30 | (0.69, 2.45) |  |
| **Did you have enough support around breastfeeding in hospital?** |  |  |  |  |  | <0.001 |  |  | 0.175 |
| No | 175 | 49 | (28.0) | 1.00 | (Ref) |  | 1.00 | (Ref) |  |
| Yes | 321 | 48 | (15.0) | 0.45 | (0.29, 0.71) |  | 0.67 | (0.37, 1.20) |  |
| **Lockdown prevented me reaching out to healthcare providers for help with issues** |  |  |  |  |  | 0.040 |  |  | 0.420 |
| No | 368 | 64 | (17.4) | 1.00 | (Ref) |  | 1.00 | (Ref) |  |
| Yes | 128 | 33 | (25.8) | 1.65 | (1.02, 2.66) |  | 1.31 | (0.68, 2.54) |  |
| **Lockdown at time of birth** |  |  |  |  |  | <0.001 |  |  | 0.021 |
| Pre-lockdown | 78 | 3 | (3.8) | 1.00 | (Ref) |  | 1.00 | (Ref) |  |
| Lockdown | 418 | 94 | (22.5) | 7.25 | (2.24, 23.53) |  | 5.02 | (1.28, 19.66) |  |
| **Have you been diagnosed with Covid-19?** |  |  |  |  |  | 0.039 |  |  | 0.038 |
| No | 471 | 88 | (18.7) | 1.00 | (Ref) |  | 1.00 | (Ref) |  |
| Yes | 25 | 9 | (36.0) | 2.45 | (1.05, 5.72) |  | 3.17 | (1.07, 9.42) |  |
| **Covid-19 has affected my ability to pay rent** |  |  |  |  |  | 0.019 |  |  | 0.304 |
| No | 425 | 77 | (18.1) | 1.00 | (Ref) |  | 1.00 | (Ref) |  |
| Yes | 65 | 20 | (30.8) | 2.01 | (1.12, 3.59) |  | 1.74 | (0.60, 5.03) |  |
| **Covid-19 has affected my ability to pay for food** |  |  |  |  |  | 0.135 |  |  | 0.033 |
| No | 431 | 81 | (18.8) | 1.00 | (Ref) |  | 1.00 | (Ref) |  |
| Yes | 59 | 16 | (27.1) | 1.61 | (0.86, 3.00) |  | 0.28 | (0.09, 0.90) |  |
| **Covid-19 has affected my ability to pay for other essentials** |  |  |  |  |  | 0.004 |  |  | 0.700 |
| No | 374 | 63 | (16.8) | 1.00 | (Ref) |  | 1.00 | (Ref) |  |
| Yes | 116 | 34 | (29.3) | 2.05 | (1.26, 3.32) |  | 1.15 | (0.57, 2.31) |  |
| OR = odds ratio; CI = confidence interval; P = p value from logistic regression model |  |  |  |  |  |  |  |  |  |

##### Mental health

Multivariable analysis showed that respondents aged between 25 and 34 years (aOR 0.25; CI 0.06 - 1.01) and aged 35 years or older (aOR 0.16; CI 0.04-0.68) were less likely to have symptoms of probable minor depression in comparison to those aged between 18 and 24 years. Those who self-identified as having predominantly negative emotions in the new parenting phase were more likely to show symptoms of probable minor depression (aOR 10.75; CI 5.41-21.37). Further factors associated with probable minor depression were struggling with breastfeeding (aOR 2.16; CI 1.36 – 3.46); lockdown preventing health care access (aOR 2.06; CI 1.20 – 3.54) and lockdown putting financial strain on families, causing issues paying for living expenses such as rent (aOR 2.58; CI 1.08 – 6.18) (Table 3).

**Table 3:** Factors associated with probable minor depression, variables with p<0.2 in the univariable analysis include in multivariable analysis.

|  |  |  |  |  |  |  |  |  |  |
| --- | --- | --- | --- | --- | --- | --- | --- | --- | --- |
|  | Study population |  |  | Unadjusted analysis |  |  | Adjusted analysis |  |  |
|  | Subjects (N) | Edinburgh scale, minor depression N | Edinburgh scale, minor depression (%) | OR | 95% CI | P | OR | 95% CI | P |
| All subjects | 496 | 314 | (63.3) |  |  |  |  |  |  |
| Study variables |  |  |  |  |  |  |  |  |  |
| Age (years) |  |  |  |  |  | 0.007 |  |  | 0.023 |
| 18-24 | 21 | 18 | (85.7) | 1.00 | (Ref) |  | 1.00 | (Ref) |  |
| 25-24 | 326 | 215 | (66.0) | 0.32 | (0.09, 1.12) |  | 0.25 | (0.06, 1.01) |  |
| 35 and above | 149 | 81 | (54.4) | 0.20 | (0.06, 0.70) |  | 0.16 | (0.04, 0.68) |  |
| Highest education level completed |  |  |  |  |  | 0.048 |  |  | 0.729 |
| High school | 105 | 77 | (73.3) | 1.00 | (Ref) |  | 1.00 | (Ref) |  |
| Undergrad | 183 | 114 | (62.3) | 0.60 | (0.36, 1.02) |  | 0.79 | (0.42, 1.51) |  |
| Postgrad | 208 | 123 | (59.1) | 0.53 | (0.31, 0.88) |  | 0.78 | (0.41, 1.48) |  |
| Predominant birth emotion negative |  |  |  |  |  | <0.001 |  |  | 0.151 |
| Positive | 399 | 231 | (57.9) | 1.00 | (Ref) |  | 1.00 | (Ref) |  |
| Negative | 97 | 83 | (85.6) | 4.31 | (2.37, 7.86) |  | 1.70 | (0.82, 3.52) |  |
| Predominant negative emotions during the new parenting phase |  |  |  |  |  | <0.001 |  |  | <0.001 |
| Positive | 345 | 174 | (50.4) | 1.00 | (Ref) |  | 1.00 | (Ref) |  |
| Negative | 151 | 140 | (92.7) | 12.51 | (6.54, 23.94) |  | 10.75 | (5.41, 21.37) |  |
| Actual birth method |  |  |  |  |  | 0.118 |  |  | 0.133 |
| C-section | 311 | 205 | (65.9) | 1.00 | (Ref) |  | 1.00 | (Ref) |  |
| Vaginal | 185 | 109 | (58.9) | 0.74 | (0.51, 1.08) |  | 0.69 | (0.43, 1.12) |  |
| Was your baby placed on you skin-to skin soon after birth? |  |  |  |  |  | 0.036 |  |  | 0.979 |
| No | 116 | 83 | (71.6) | 1.00 | (Ref) |  | 1.00 | (Ref) |  |
| Yes | 380 | 231 | (60.8) | 0.62 | (0.39, 0.97) |  | 0.99 | (0.56, 1.76) |  |
| Were you discharged from hospital after the birth earlier than expected because |  |  |  |  |  | 0.086 |  |  | 0.149 |
| No | 359 | 219 | (61.0) | 1.00 | (Ref) |  | 1.00 | (Ref) |  |
| Yes | 137 | 95 | (69.3) | 1.45 | (0.95, 2.20) |  | 1.45 | (0.87, 2.42) |  |
| Struggled with breastfeeding |  |  |  |  |  | <0.001 |  |  | 0.001 |
| No | 221 | 122 | (55.2) | 1.00 | (Ref) |  | 1.00 | (Ref) |  |
| Yes | 275 | 192 | (69.8) | 1.88 | (1.30, 2.72) |  | 2.16 | (1.36, 3.46) |  |
| **Did you have enough support around breastfeeding in hospital?** |  |  |  |  |  | 0.110 |  |  | 0.713 |
| No | 175 | 119 | (68.0) | 1.00 | (Ref) |  | 1.00 | (Ref) |  |
| Yes | 321 | 195 | (60.7) | 0.73 | (0.49, 1.07) |  | 1.10 | (0.67, 1.80) |  |
| **Did you access any mental health support services after your 2020 birth?** |  |  |  |  |  | 0.014 |  |  | 0.111 |
| No | 427 | 261 | (61.1) | 1.00 | (Ref) |  | 1.00 | (Ref) |  |
| Yes | 69 | 53 | (76.8) | 2.11 | (1.17, 3.81) |  | 1.74 | (0.88, 3.44) |  |
| **Lockdown prevented me reaching out to healthcare providers for help with issues** |  |  |  |  |  | <0.001 |  |  | 0.009 |
| No | 368 | 213 | (57.9) | 1.00 | (Ref) |  | 1.00 | (Ref) |  |
| Yes | 128 | 101 | (78.9) | 2.72 | (1.70, 4.37) |  | 2.06 | (1.20, 3.54) |  |
| **Have you been diagnosed with Covid-19?** |  |  |  |  |  | 0.084 |  |  | 0.226 |
| No | 471 | 294 | (62.4) | 1.00 | (Ref) |  | 1.00 | (Ref) |  |
| Yes | 25 | 20 | (80.0) | 2.41 | (0.89, 6.53) |  | 1.98 | (0.66, 5.95) |  |
| **Covid-19 has affected my ability to pay rent** |  |  |  |  |  | 0.004 |  |  | 0.033 |
| No | 425 | 259 | (60.9) | 1.00 | (Ref) |  | 1.00 | (Ref) |  |
| Yes | 65 | 52 | (80.0) | 2.56 | (1.35, 4.85) |  | 2.58 | (1.08, 6.18) |  |
| **Covid-19 has affected my ability to pay for food** |  |  |  |  |  | 0.007 |  |  | 0.582 |
| No | 431 | 264 | (61.3) | 1.00 | (Ref) |  | 1.00 | (Ref) |  |
| Yes | 59 | 47 | (79.7) | 2.48 | (1.28, 4.81) |  | 0.75 | (0.27, 2.09) |  |
| **Covid-19 has affected my ability to pay for other essentials** |  |  |  |  |  | <0.001 |  |  | 0.063 |
| No | 374 | 221 | (59.1) | 1.00 | (Ref) |  | 1.00 | (Ref) |  |
| Yes | 116 | 90 | (77.6) | 2.40 | (1.48, 3.88) |  | 1.79 | (0.97, 3.31) |  |
| OR = odds ratio; CI = confidence interval; P = p value from logistic regression model |  |  |  |  |  |  |  |  |  |

#### Support

Very few mothers (n=69; 14%) reported accessing professional support for mental health after their birth. However, several reported accessing other forms of support via social media (n=328; 66%), websites (n=309; 62%) and on WhatsApp moms groups (n=226; 46%). Most (n=474; 96%) moms are able to access a reliable internet connection (Table 4).

**Table 4:** Support accessed by mothers after birth in 2020.

|  |  | **N=496** | **%** |
| --- | --- | --- | --- |
| Did you access any mental health support services after your 2020 birth? | Yes | 69 | 13.9% |
|  | No | 427 | 86.1% |
| Did you find any of these resources helpful during this time? | Social media | 328 | 66.1% |
|  | Websites | 309 | 62.3% |
|  | WhatsApp moms groups | 226 | 45.6% |
|  | Blogs | 173 | 34.9% |
|  | Other | 46 | 9.3% |
| Did you have a reliable internet connection to be able to access support online? | Yes | 474 | 95.6% |
|  | No | 22 | 4.4% |
| Were internet data charges too expensive for you to be able to use the internet to access support? | Yes | 55 | 11.1% |
|  | No | 441 | 88.9% |

### Males

The median age of male respondents (n=24) was 37 years (IQR 29, 39). Male respondents mainly reported having the preferred method of birth (n=15; 63%); feeling positive about the birth experience (n=23; 95%); and that the birth did not affect their parenting (n=17; 71%). The Covid-19 pandemic and resultant lockdown affected 30% (n=7) of fathers birth experiences, mainly due to the father not being able to visit the mother and child in hospital after the birth (n=5; 21%) and because the pandemic led to a change in expected birth plan (n=4; 17%). Twenty-five percent of (n=6) of fathers showed symptoms of major depression. The most widely used forms of online support for fathers are websites (n=20; 83%) and social media (n=11; 46%) (Table 5).

**Table 5:**
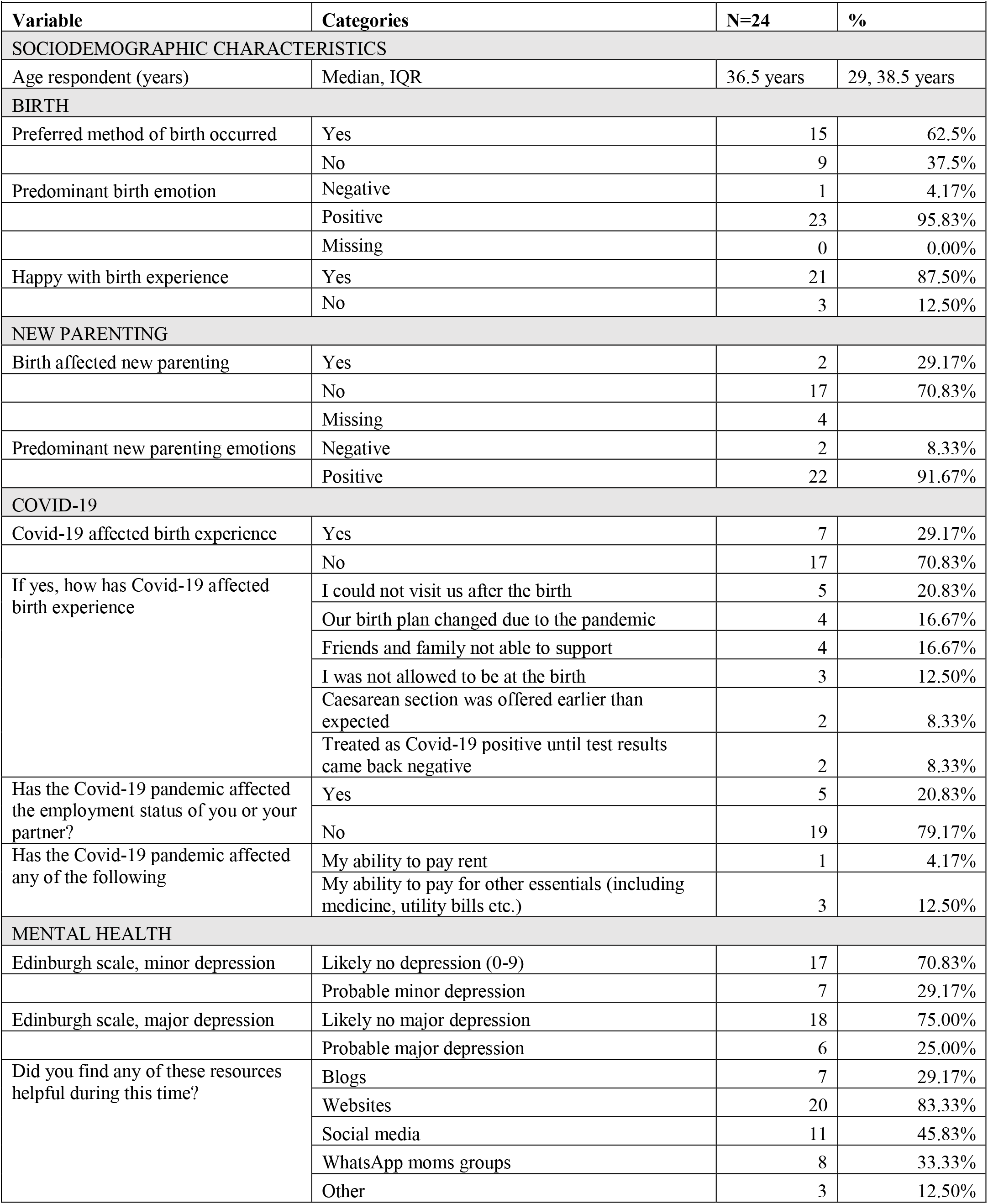
Fathers of babies born during 2020, birth and new parenting experiences.

### Thematic analysis

Thematic content analysis of open responses indicated five core themes that confirm and deepen the quantitative findings of birth and new parenting experiences under COVID-19.

#### Social exclusion and isolation leading to increased stress and anxiety

Overwhelmingly the most common theme (n=169), new parents reported strong feelings of social exclusion and isolation, particularly during the birthing experience that impacted on their levels of stress and anxiety as new parents. The lack of social support due to the forced absence of a partner, close family members and friends were the most cited reasons for feelings of social isolation and loneliness.

> *“Not being able to be with my parents. My mom for help and of course my sister. I never envisioned not getting help, love and support. The celebration I envisioned was taken away.”*

Interestingly, for some parents the absence of outside family and friends was seen as a positive opportunity for the new family to bond without interruption. Although, this was the case for a significant minority of respondents and typically when at least one significant other was allowed to be present:

> *“No visitors allowed at the hospital. I saw this as a positive so my husband and I could bond with new-born and become a little family unit”*

#### Rapidly modified health services impact experiences of quality and quantity of care

The second most commonly reported theme (n=88) indicated how rapidly modified health services impacted new parents’ experiences of both the quality and quantity of care received during lockdown. New COVID-19 testing protocols, shorter hospital stays and reduced availability of expected services and appointments (e.g. lactation consultants, midwives, gynaecologists) reduced the quantity of care reported by new parents. While, unclear policies and procedures, conflicting medical advice, unexpected medical procedures and reduced availability and attentiveness of medical personnel impacted perceptions on the quality of care received. These issues are summarised by one new mother who reported her hospital experience during lockdown:

> *“The Covid test was done 30 minutes after being booked into the hospital, but the test was never done. So when my baby was born 16 hours later, I wasn’t allowed near her as my test results were not back yet. I only held my baby 7 hours after she was born - she was given formula without my consent. Dad wasn’t allowed in, so he only saw her when we were discharged. I was also served food and drink in take away containers as my test results were not back yet. Only after tests came back negative, were I given food in a plate.”*

#### Anxiety and the virus: increasing the “mental load of motherhood”

Feelings of fear and anxiety linked to COVID-19 permeated the pre-, peri- and post-natal experiences reported by new mothers (n=64). Anxiety was specifically described due to fear of infection, the implications of COVID-19 regulations on access to healthcare and social support during birth, managing work and additional childcare responsibilities, and the social reintegration for both family and baby once lockdown eased. For some, the anxiety of COVID-19 prevented them from seeking the healthcare and support they needed as new parents:

> *“I’m scared to take my baby anywhere because of Covid-19. I should have done a breastfeeding course prior to delivery but I did not because I was too scared of contracting the virus.”*

While others reported the significant impact of COVID-19 and the ensuing lockdown on their mental health and wellbeing:

> *“Increased mental load of motherhood. Working from home with pre-schooler and baby. Struggle with post-partum anxiety and depression.”*

These findings highlight quantitative results that emphasize the need for increased access to mental health support services for new parents, especially under COVID-19 conditions.

#### The impact of changing work and economic circumstances

Changing work and economic circumstances due to COVID-19 lockdown added additional stress to birth and new parenting experiences (n=51). The economic impact of lockdown introduced higher levels of uncertainty with reports of reduced income, delayed UIF payments, retrenchments and job losses. Combined with reduced access to childcare and the challenge of looking for new work while also adjusting to the arrival of a new baby, many parents were forced to make significant changes to their living circumstances:

> *“Due to my husband losing his income we are financially in a crisis (even still). We had to move when my baby was just 2 weeks old due to COVID. We cannot afford any help from nanny or schooling. I had to go back to work sooner in order to receive a salary to keep us barely afloat”*

#### A positive birth experience can support a positive transition to parenthood

Despite the rapid changes required by COVID-19 conditions, women were more likely to report a positive transition to parenting if their birthing experience had gone according to plan (n=60):

> *“I was so happy my delivery went according to plan. I felt empowered and therefore I think I went into those first few weeks much more confidently.”*

Together with feelings of empowerment and the ability to cope, a positive, supportive birthing experience was also linked to new mothers’ experiences of bonding with their babies:

> *“The calm birth set the scene for a calm baby and mom, and ensured a proper bond between us too.”*

These findings, although not universal, correspond with the overall positive emotions (80.4%) reported by new mothers in this study. They also highlight the importance of positive birthing experiences as well as the resilience of new mothers giving birth under unique Covid-19 conditions.

## Discussion

Our study showed that the Covid-19 lockdown impacted the birth and new parenting experiences of parents whose children were born in 2020. Parents with children born under lockdown were less likely to have their preferred choice of birth method, had worse self-reported birth experiences, were less likely to have skin-to-skin contact with their babies after birth and were frequently unable to have someone at their birth that they had wanted to be there. Health services were modified due to the pandemic and this impacted new parents’ experiences of both the quality and quantity of care received. Despite these challenges, parents reported positive birth and new parenting experiences, showing resilience.

Our results were similar to two studies, one from the United Kingdom [8] and the other from Italy [13], which showed that the Covid-19 lockdown led to distress for parents who reported social isolation as a key challenge. The United Kingdom study also reported that lockdown led to shorter hospital stays, changes to the delivery plans, a lack of breastfeeding support, challenges accessing healthcare, limited social support and mental health difficulties [8], findings corroborated by our study.

Our findings on the prominence of probable depression (63% mothers, 29% fathers), are higher than the usual in this context (33%), which are higher than worldwide pre-Covid-19 estimates (13%) [5]. This finding is mirrored by Canadian study which found increased levels of depression amoungst new fathers [14] and a report from an Australian postnatal support hotline that calls increased 75% during the first week of lockdown [15], indicating an increase in new parents requiring mental health support. Our study showed that mental health issues affected many respondents who were younger, had negative emotions about their birth, struggled with breastfeeding, could not access health care and had lockdown related financial concerns. Parents need support to overcome these challenges. Other studies have shown that digital methods of care provision during the post-partum period are effective [16]. These forms of care are reported in our study to be acceptable (specifically social media, websites and WhatsApp moms groups) and should form the basis of interventions for this population.

Mothers in our cohort reported that lockdown restrictions made the new parenting phase more challenging. In particular there was the ban on the sale of baby products which was introduced with Level 5 lockdown and only overturned after the restriction was ruled unconstitutional in early April 2020 [17]. A further challenge was the registration of babies, which by law should happen within 30 days of their birth. Prior to lockdown, registrations took place within hospitals, this was cancelled during lockdown, as were many services at Home Affairs, making registration difficult. The ban on exercise, which was a feature of several lockdown levels, was noted by respondents as something that impact their postpartum quality of life. Exercise in the postpartum period has been linked to several positive changes for the mother including improved psychosocial well-being in other settings [18]. Future lockdown restriction implementers and policy makers should take these challenges into consideration. Baby products should always be included in essential items lists, and be available for sale in shops and online; plans should be made to assist new parents to register their babies in a timely manner and outdoor exercise with precautions such as masks, should be allowed.

Not specific to the Covid-19 lockdown, but a noteworthy finding is the caesarean section rate in our study cohort (63%), a finding similar to other studies of private healthcare users in South Africa (60% [19]; 73% [20], 77% [21]). The WHO states that caesarean section are medically necessary for 10 - 15% of all births [22]. While undoubtedly lifesaving, caesarean sections can lead to severe and permanent physical complications [22, 23], increased post-traumatic stress [23], mental health issues [23] and impairments in quality of life [23] for the mother. Caesarean sections have also been reported to lead to challenges with infant-mother bonding [24], breastfeeding issues [23, 25], an increase risk of non-communicable diseases [26] and adverse effects on children’s sensory perception and neuropsychiatric development [24]. Our findings corroborate these reports and show that most mothers wanted to have a natural birth, and that a change in delivery method was directly associated with a negative birth experience which was associated with probable depression. Due to these potential issues, caesarean sections should only be used when medically necessary [22]. The problematically high caesarean section rate in South African private health care needs to be addressed. One such solution would be initiating a midwife led birthing process as opposed to the current Gynaecologist led process [15].

One of the main limitations of our study is that the sample of parents who responded to the survey do not represent the vast majority of South African citizens. The use of convenience sampling in order to rapidly recruit participants in a short window of time resulted in our study sample being mainly degreed, married, middle-to high-income earners who had their babies in private hospitals. The findings from our study can thus only be generalised to this portion of the population. Future studies need to investigate the birth and new parenting experiences in populations of lower socioeconomic status and of public sector patients. The use of convenience sampling, further limiting the generalisability of our results [27]. There is also the potential for recall bias in this study due to the differing lengths of time between the birth and completion of survey, and it is difficult to tease out the independent effects of the lockdown levels of different postpartum periods.

Covid-19 and the ensuing lockdown exacerbated many of the usual challenges of birth and new parenting. Mental health was a key challenge, along with difficulties accessing support. However, overall experiences were positive and there was a strong sense of resilience amongst parents. As the world continues to battle the Covid-19 pandemic on all fronts, efforts to support new parents during this critical phase should not be neglected.

## Data Availability

Data are available on request from

## Author statements

### Acknowledgements

The authors are grateful to the parents of the babies born during lockdown who agreed to be a part of this study.

### Author contributions

All authors were responsible for the study concept, design, data collection, analysis, and interpretation of data. All authors drafted and critically reviewed the manuscript.

## Supplementary material

**S1.**
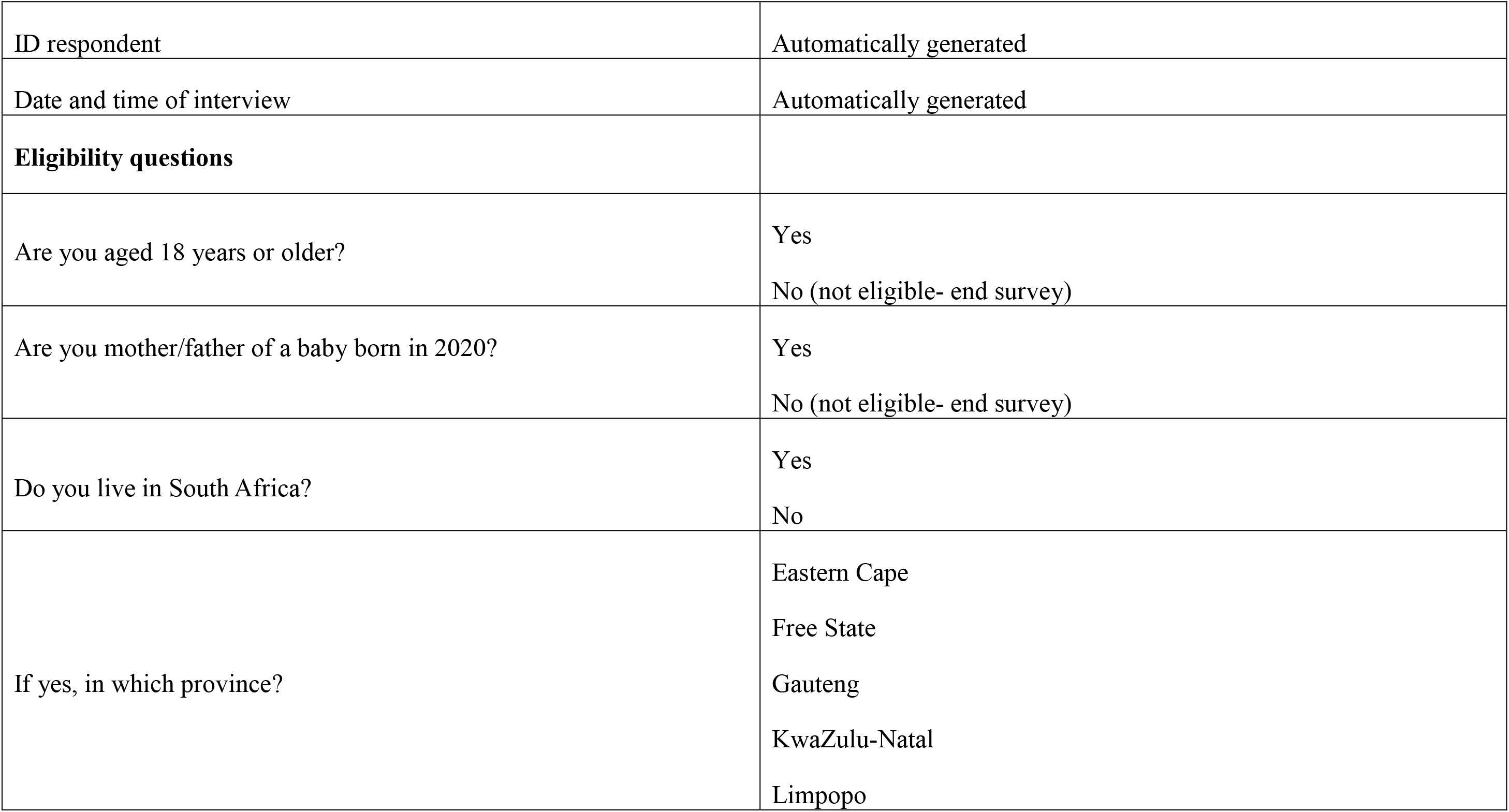
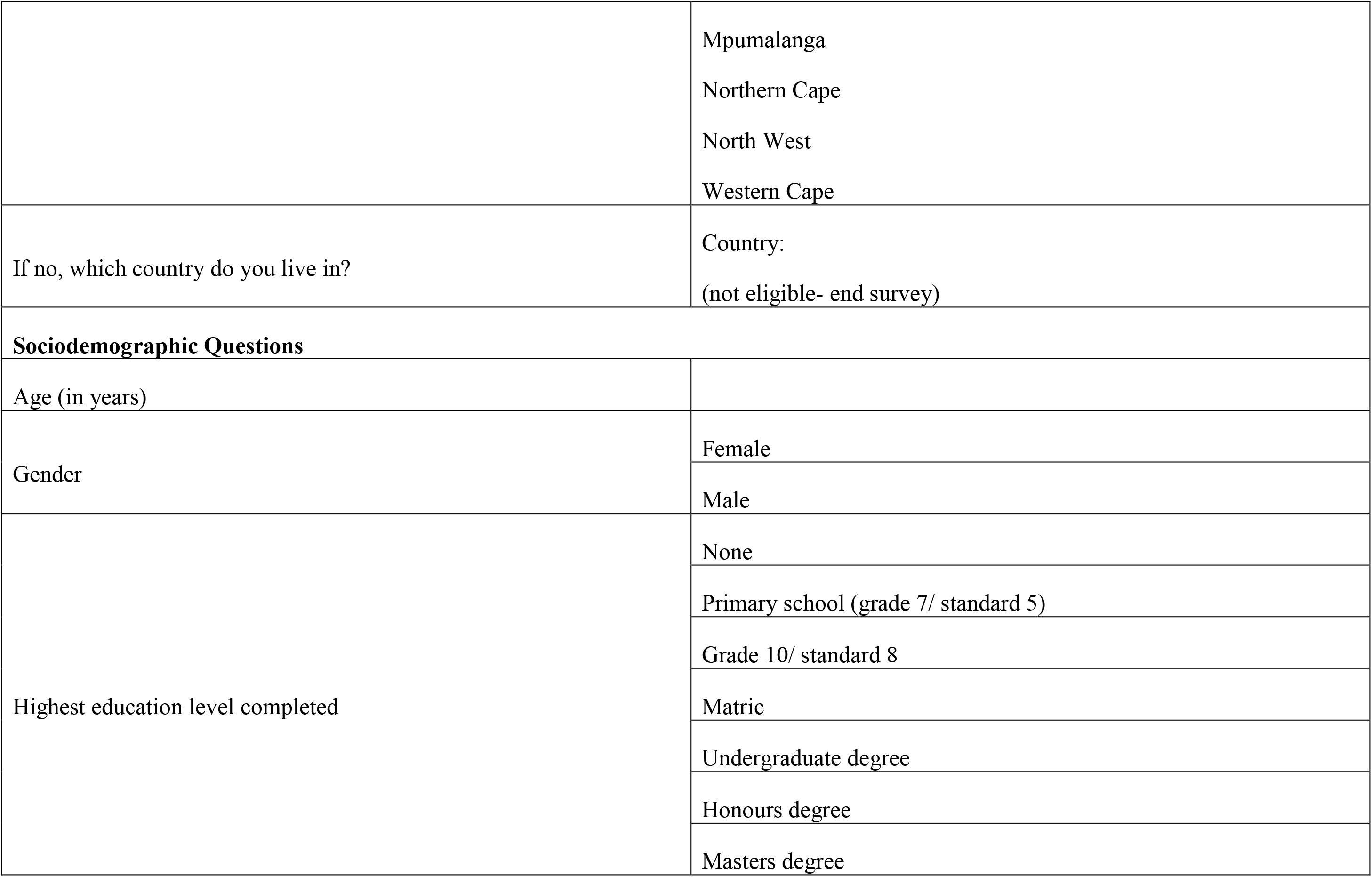
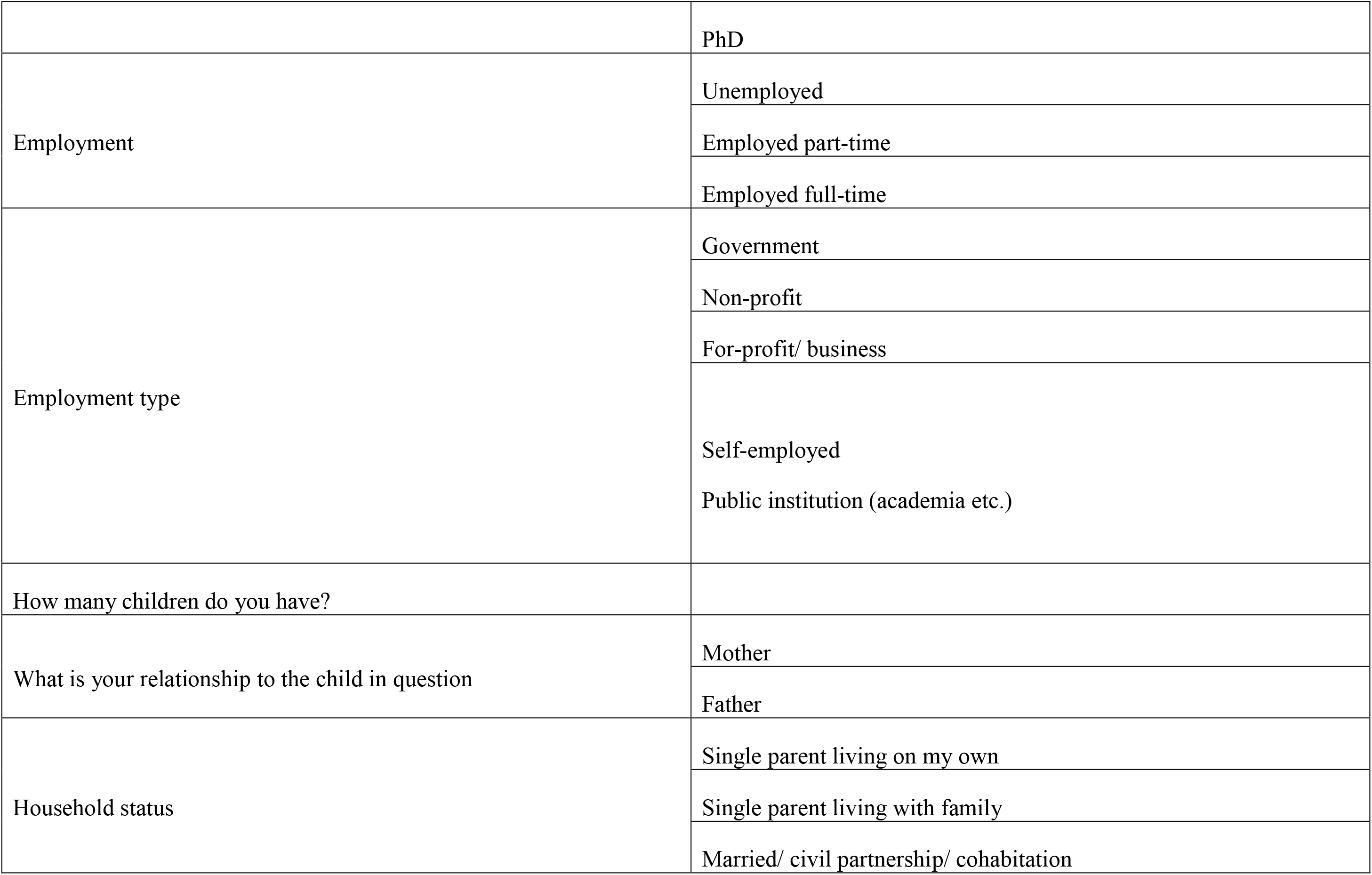
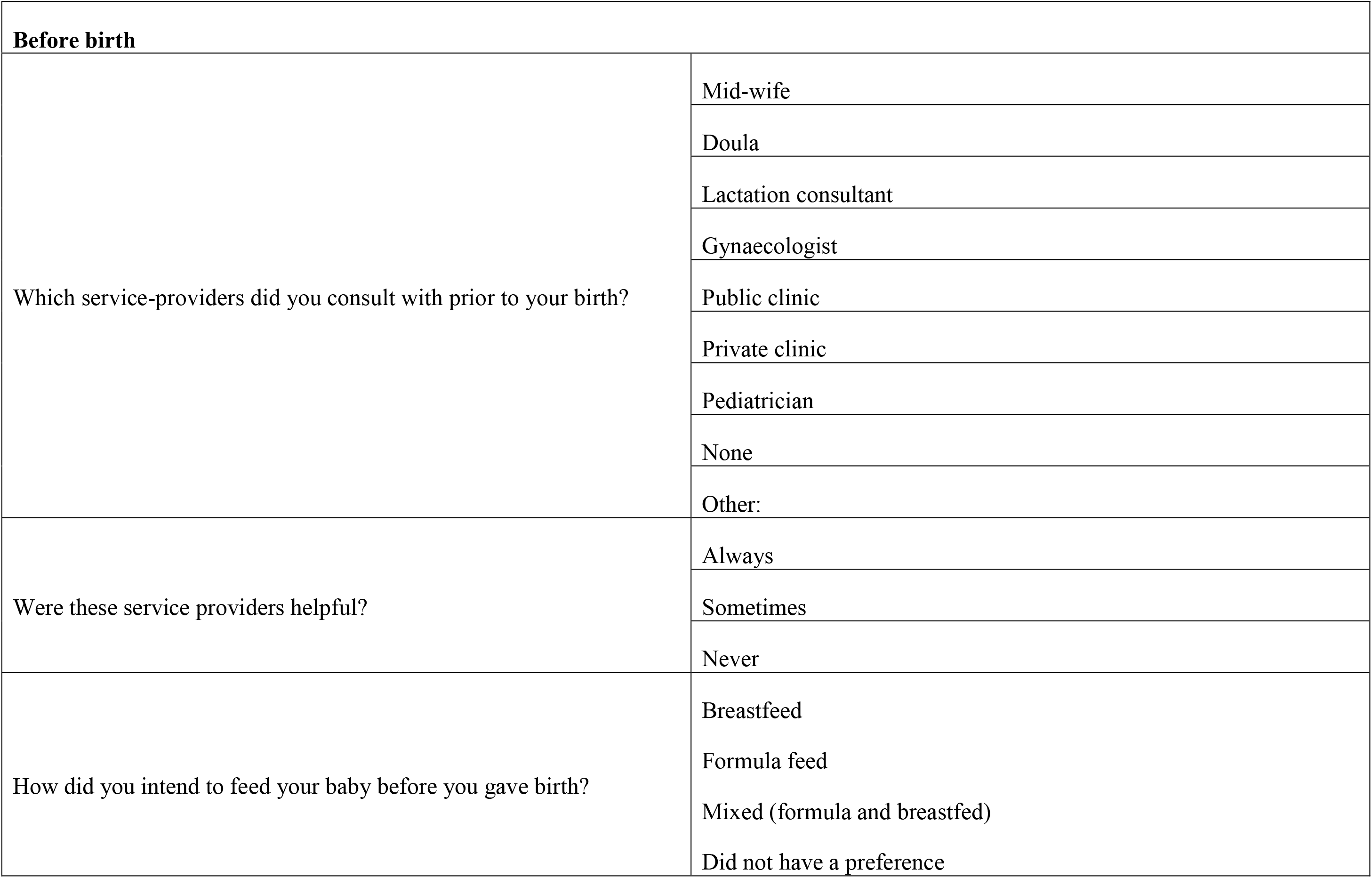
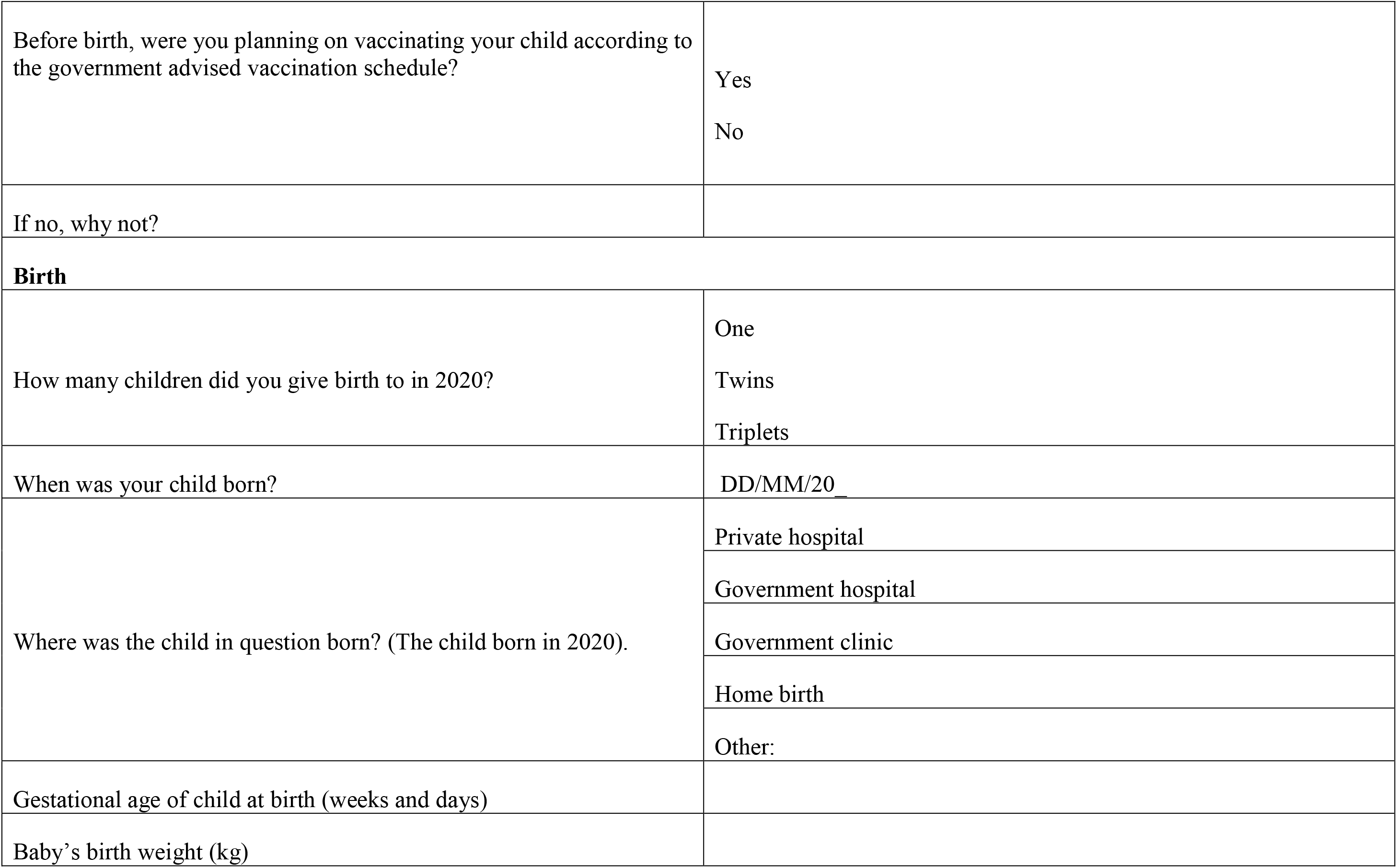
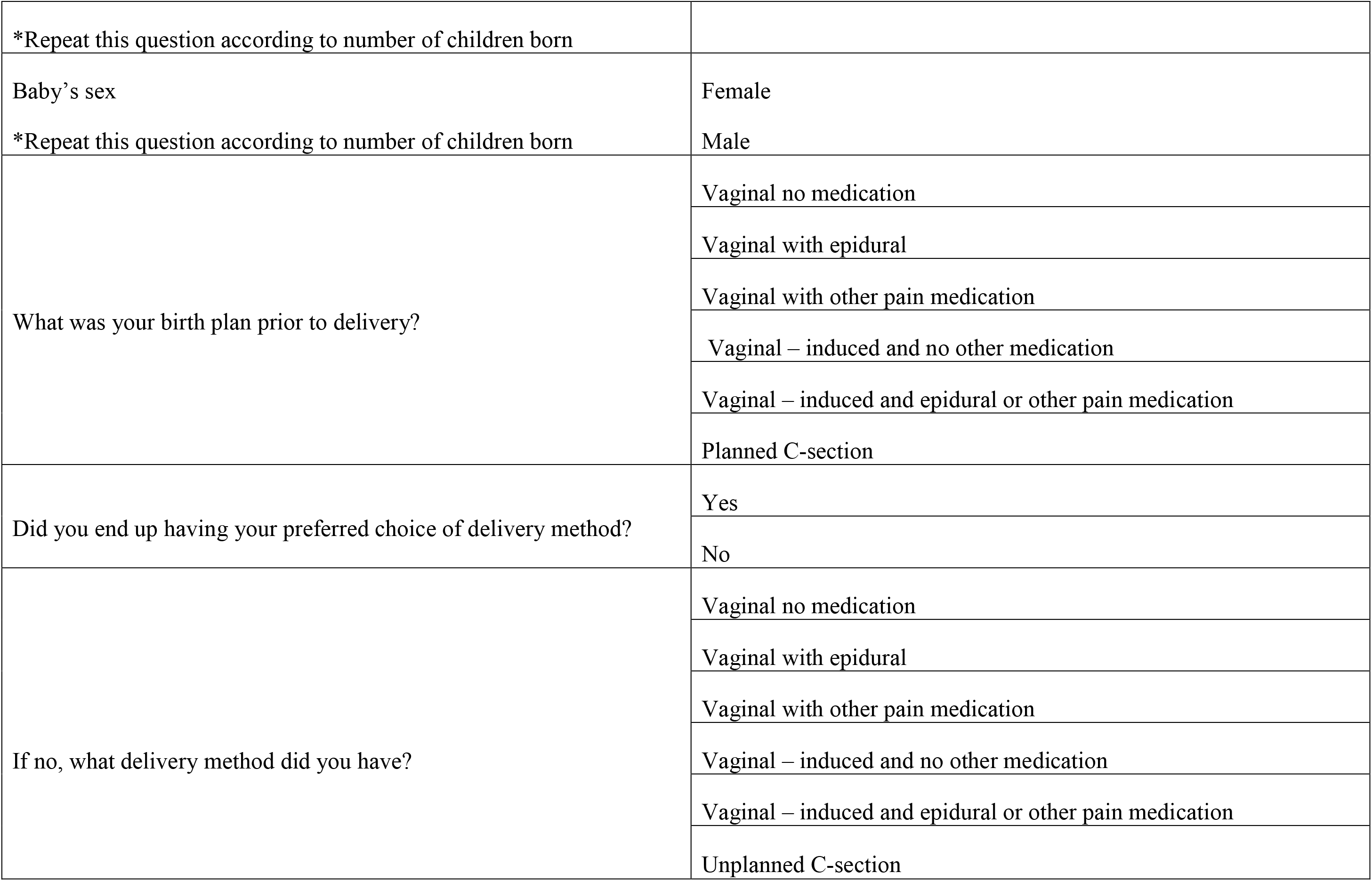
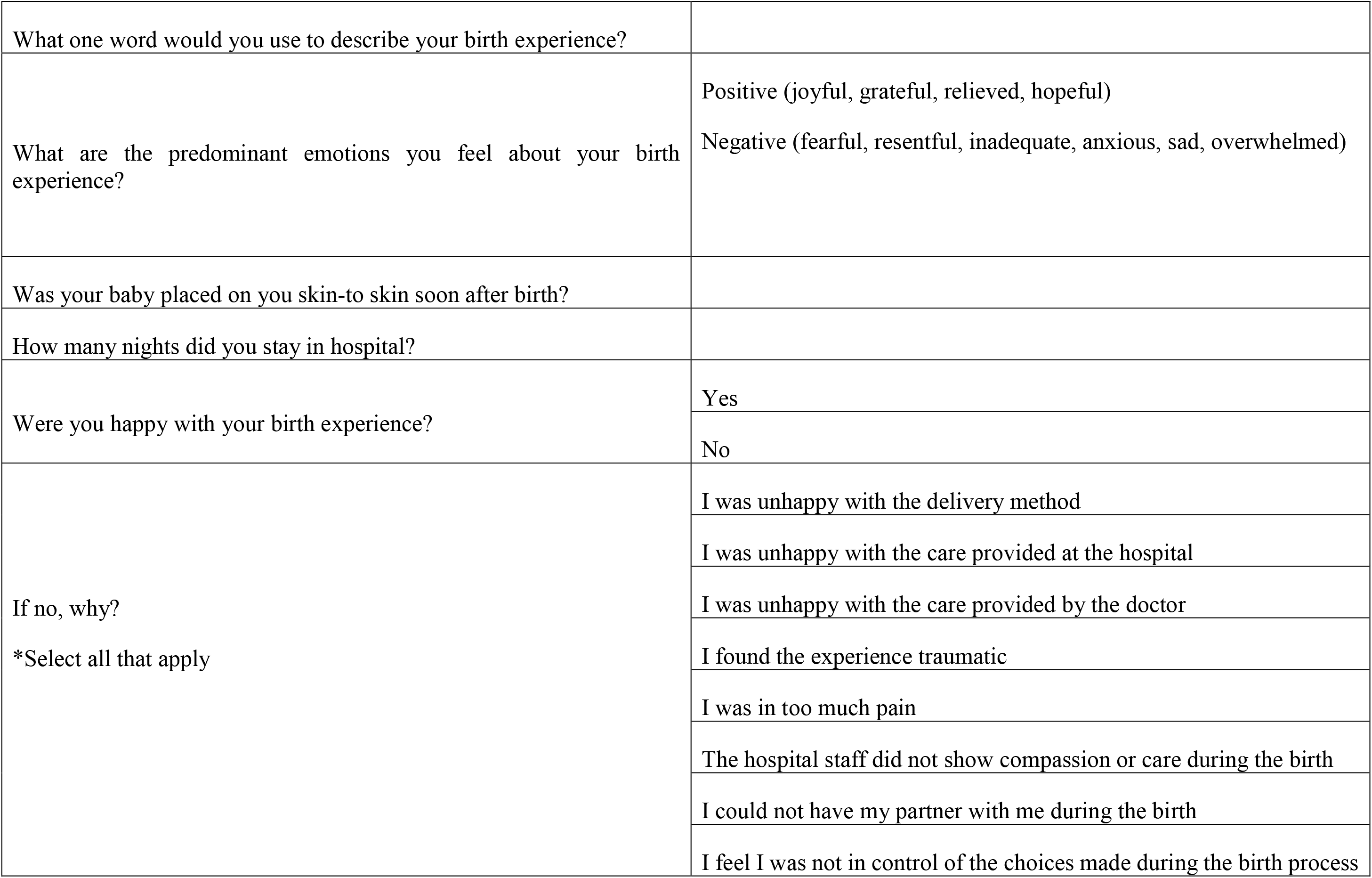
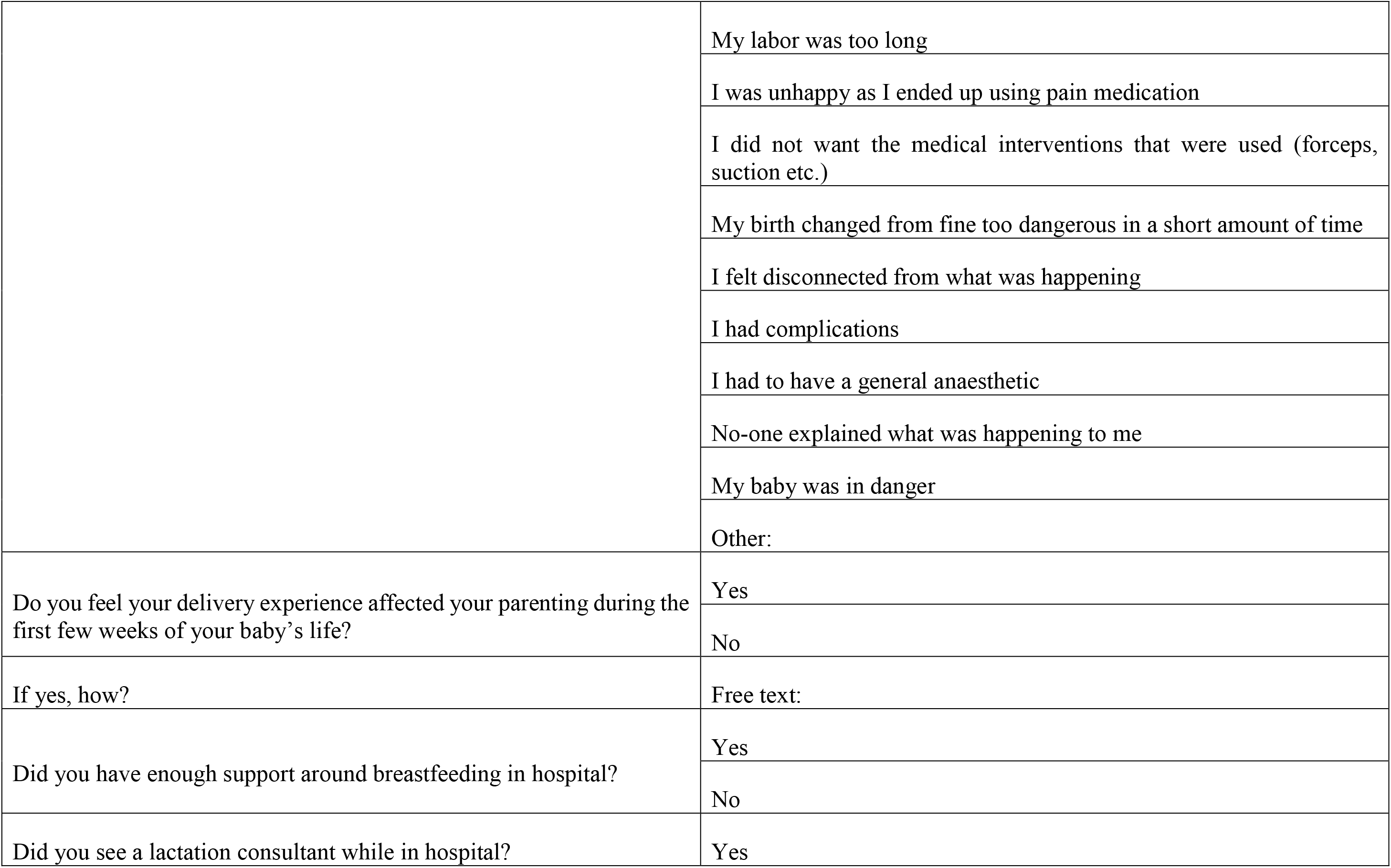
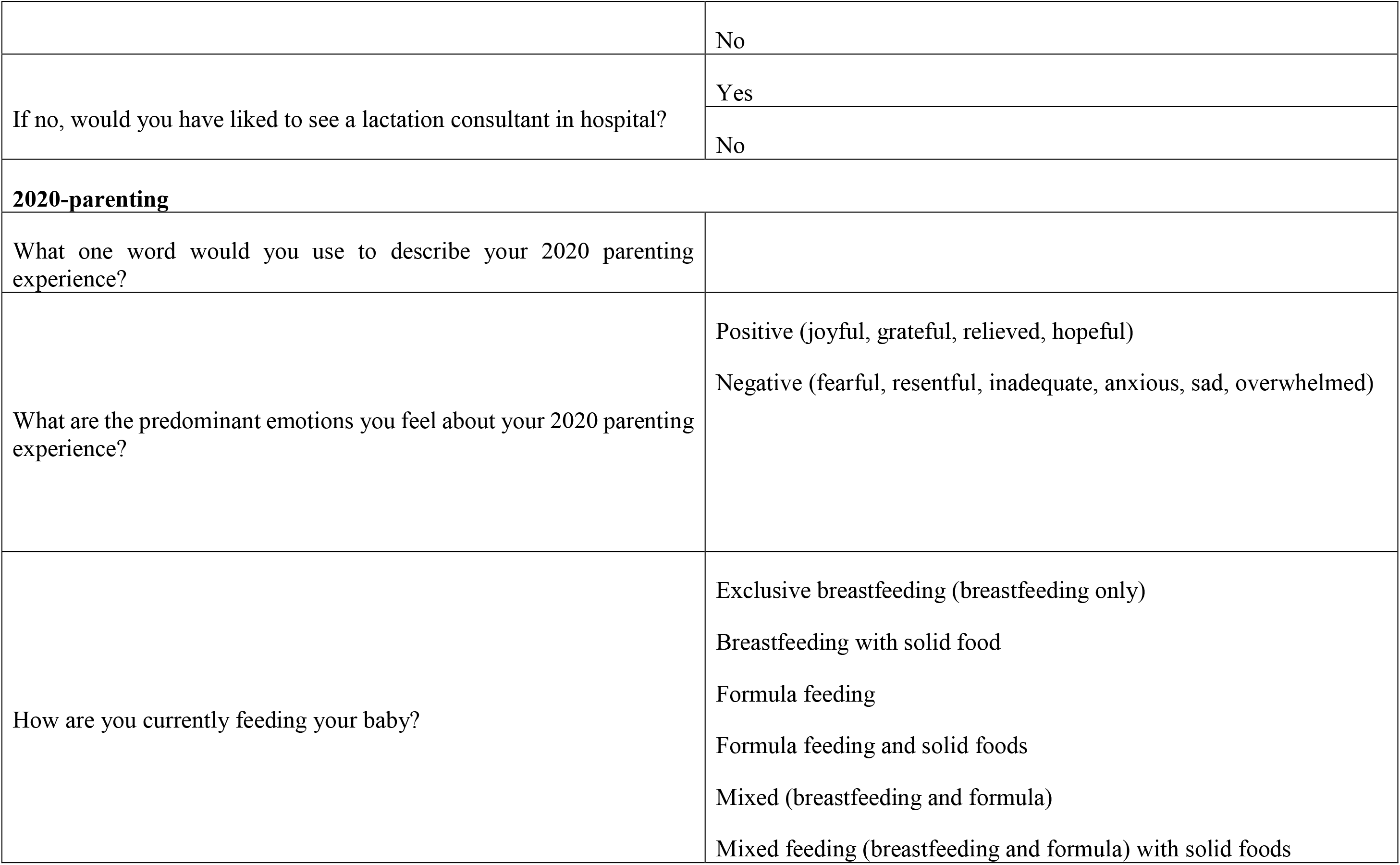
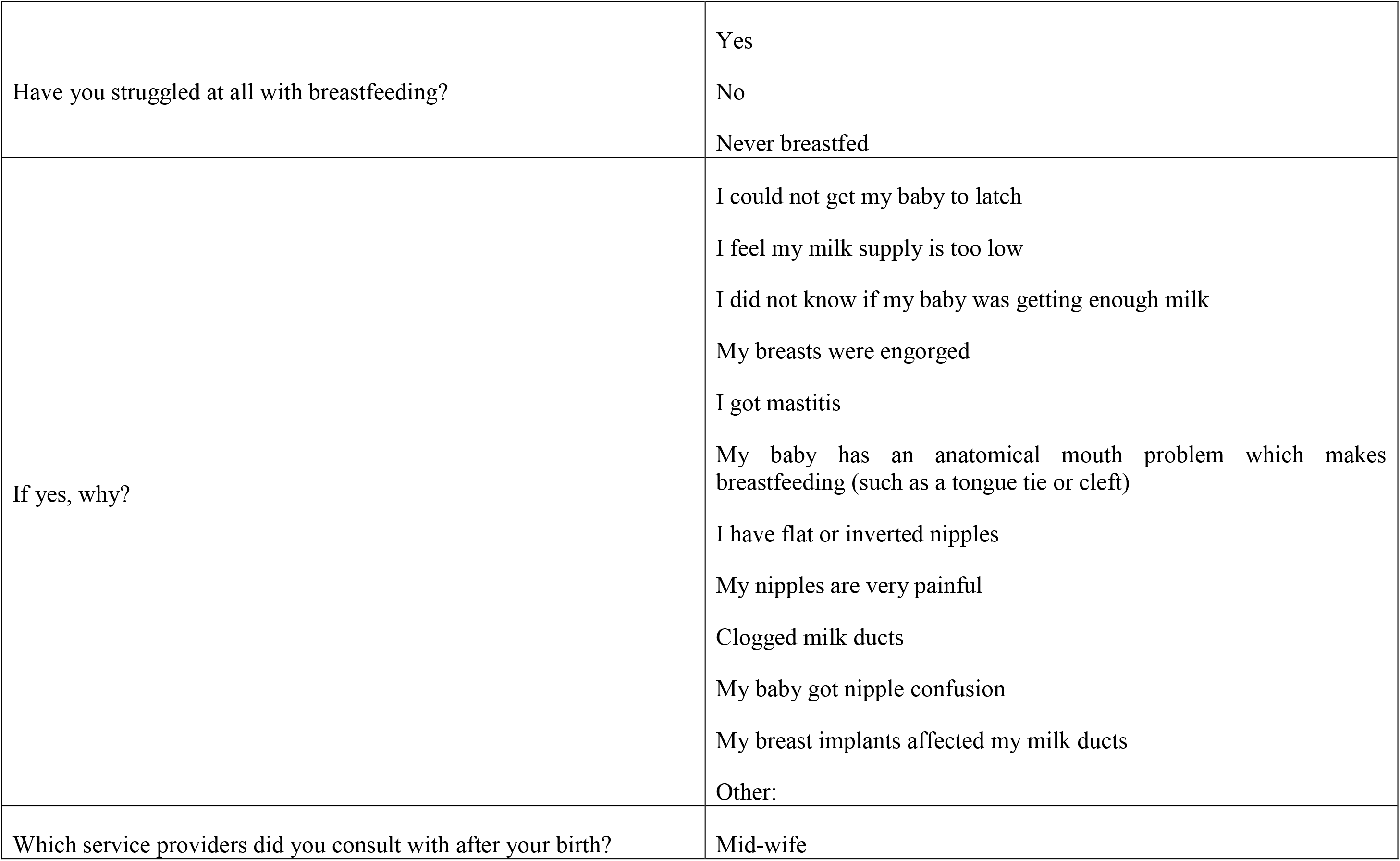
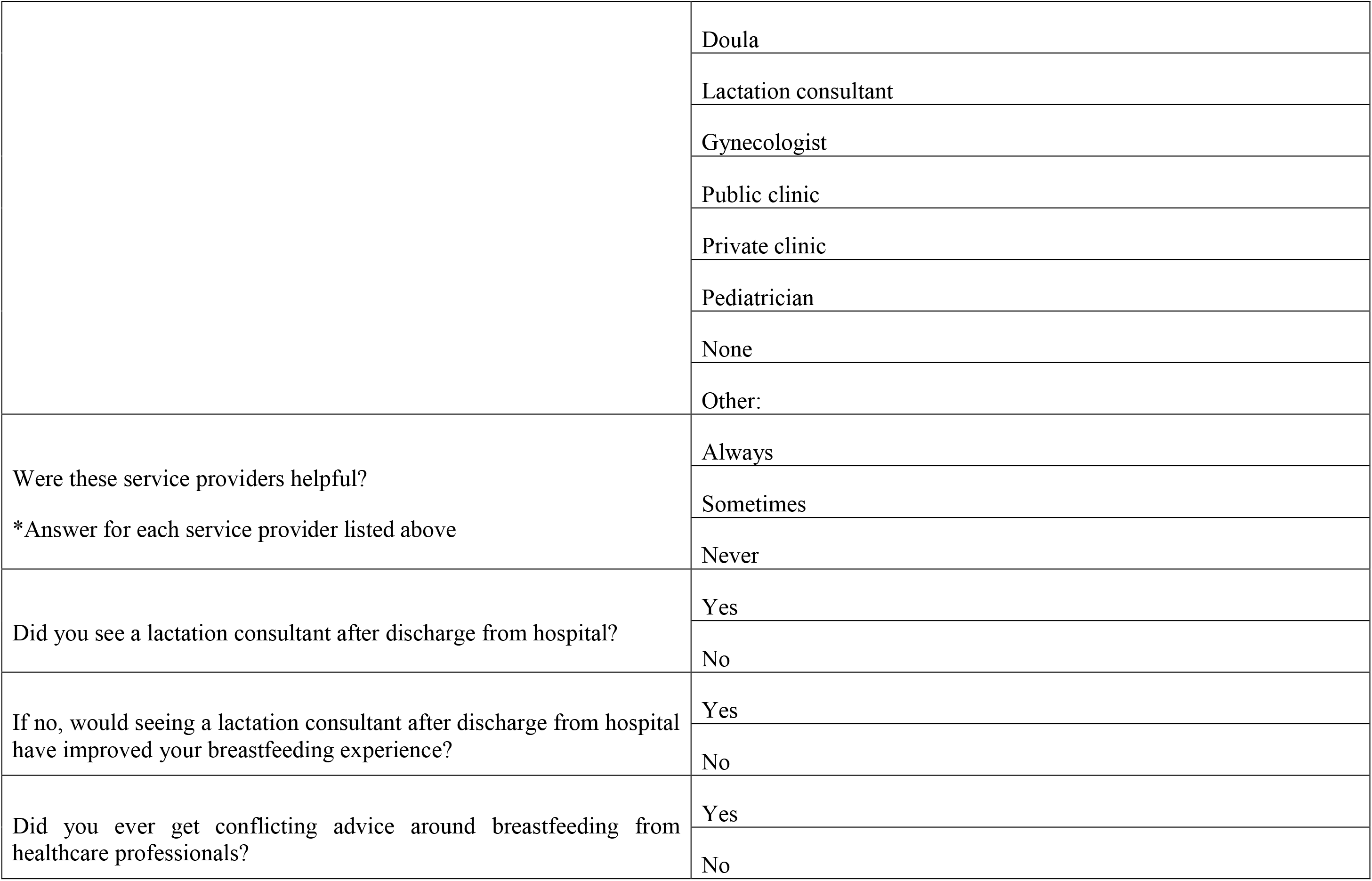
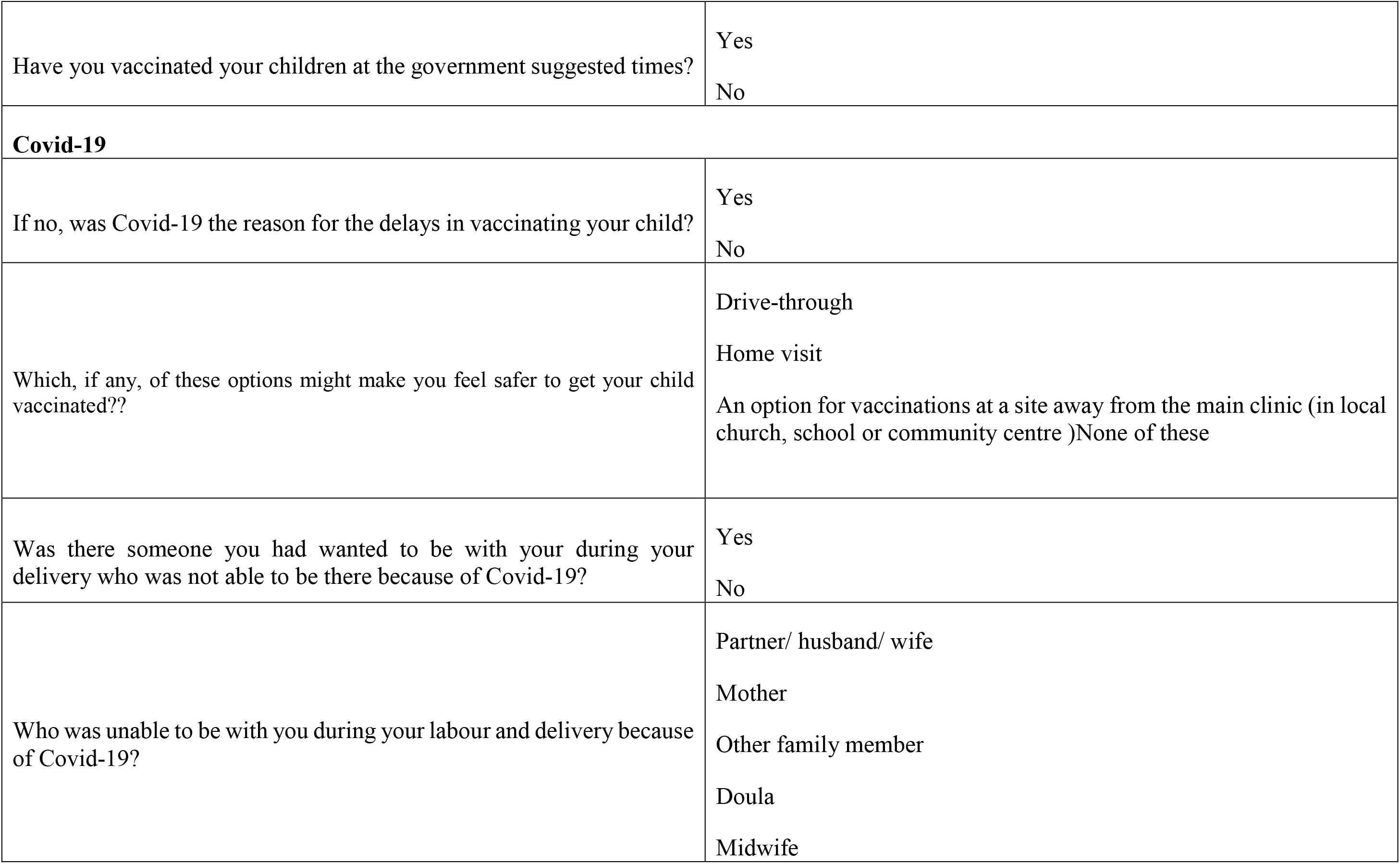
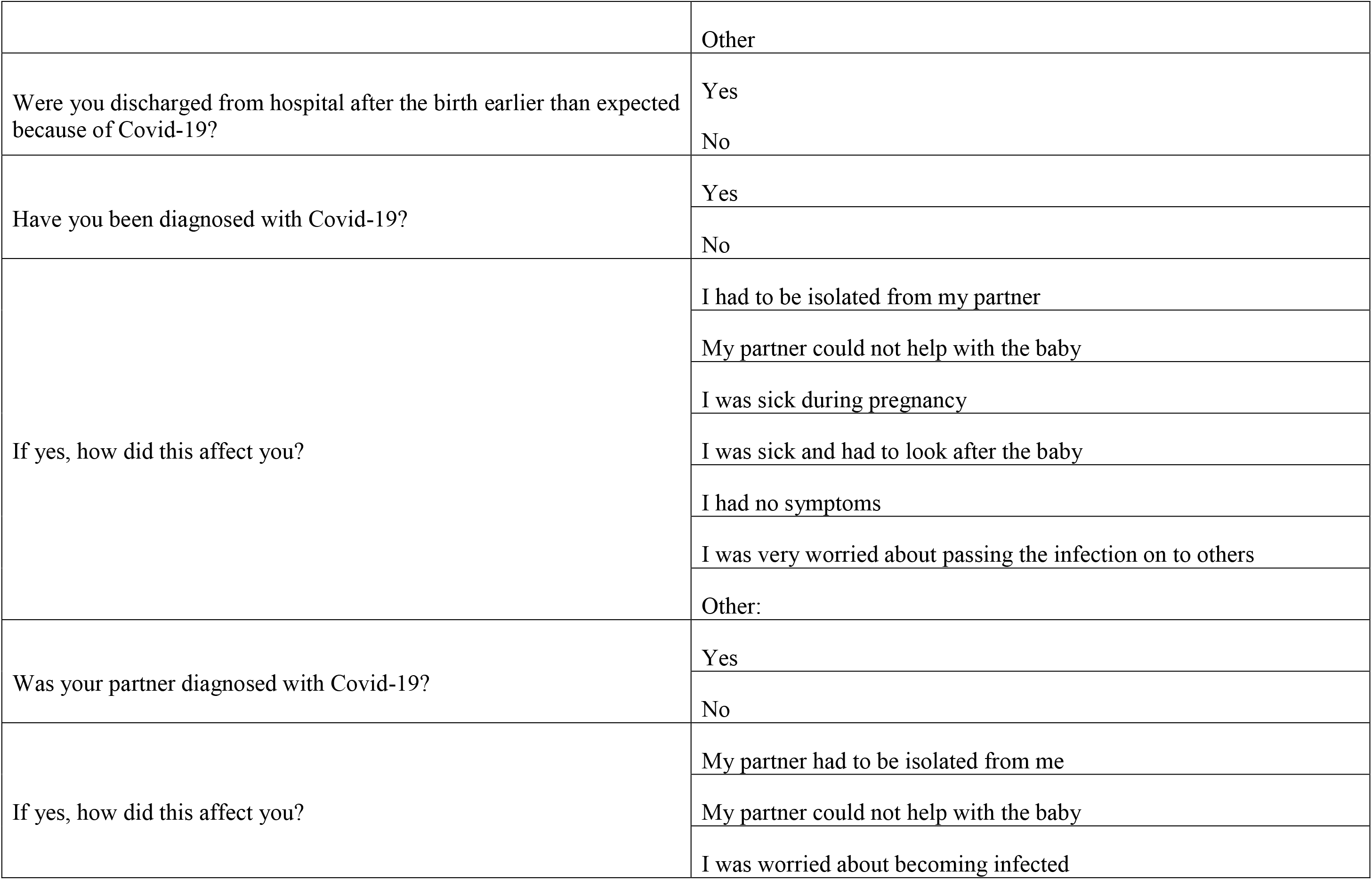
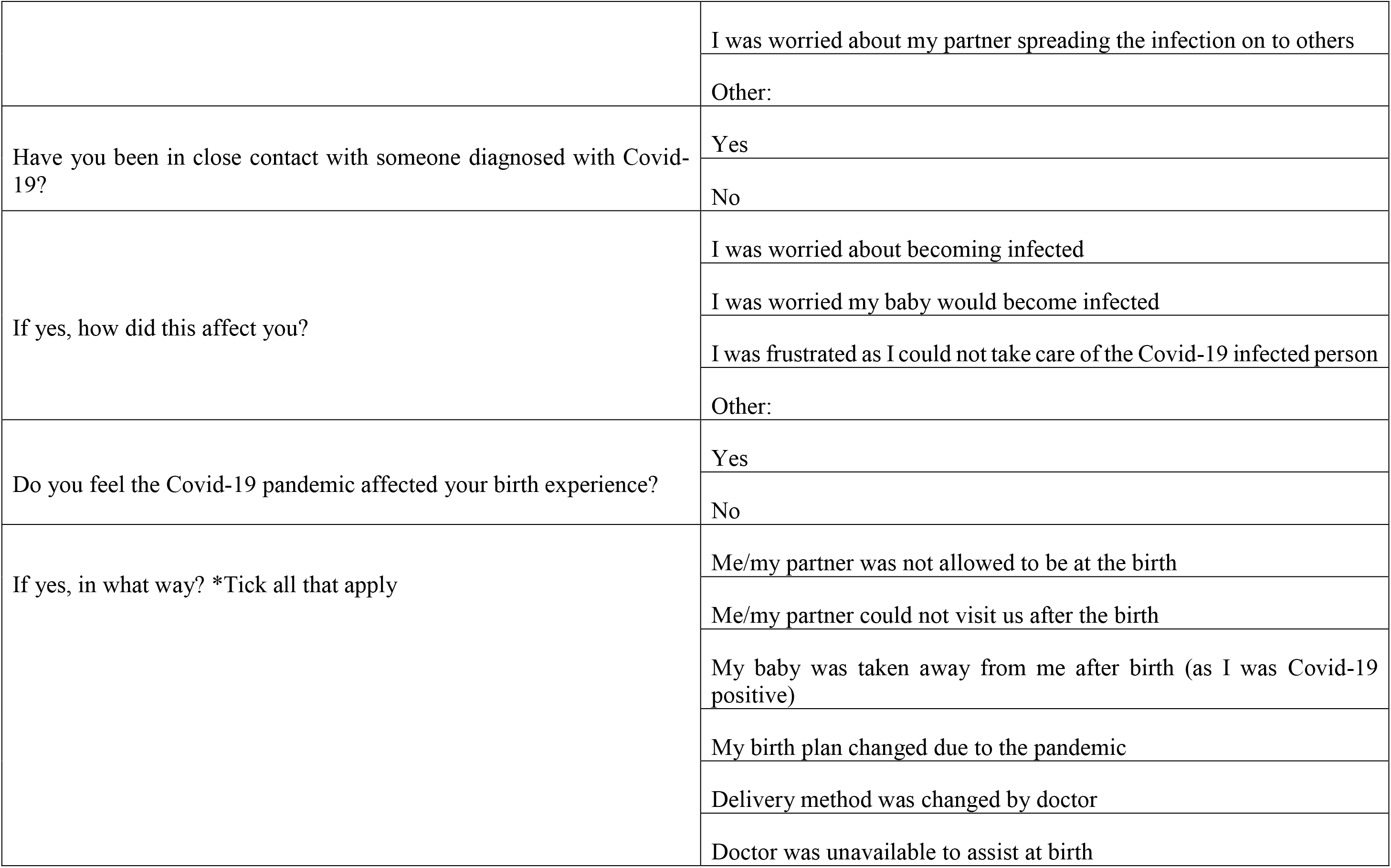
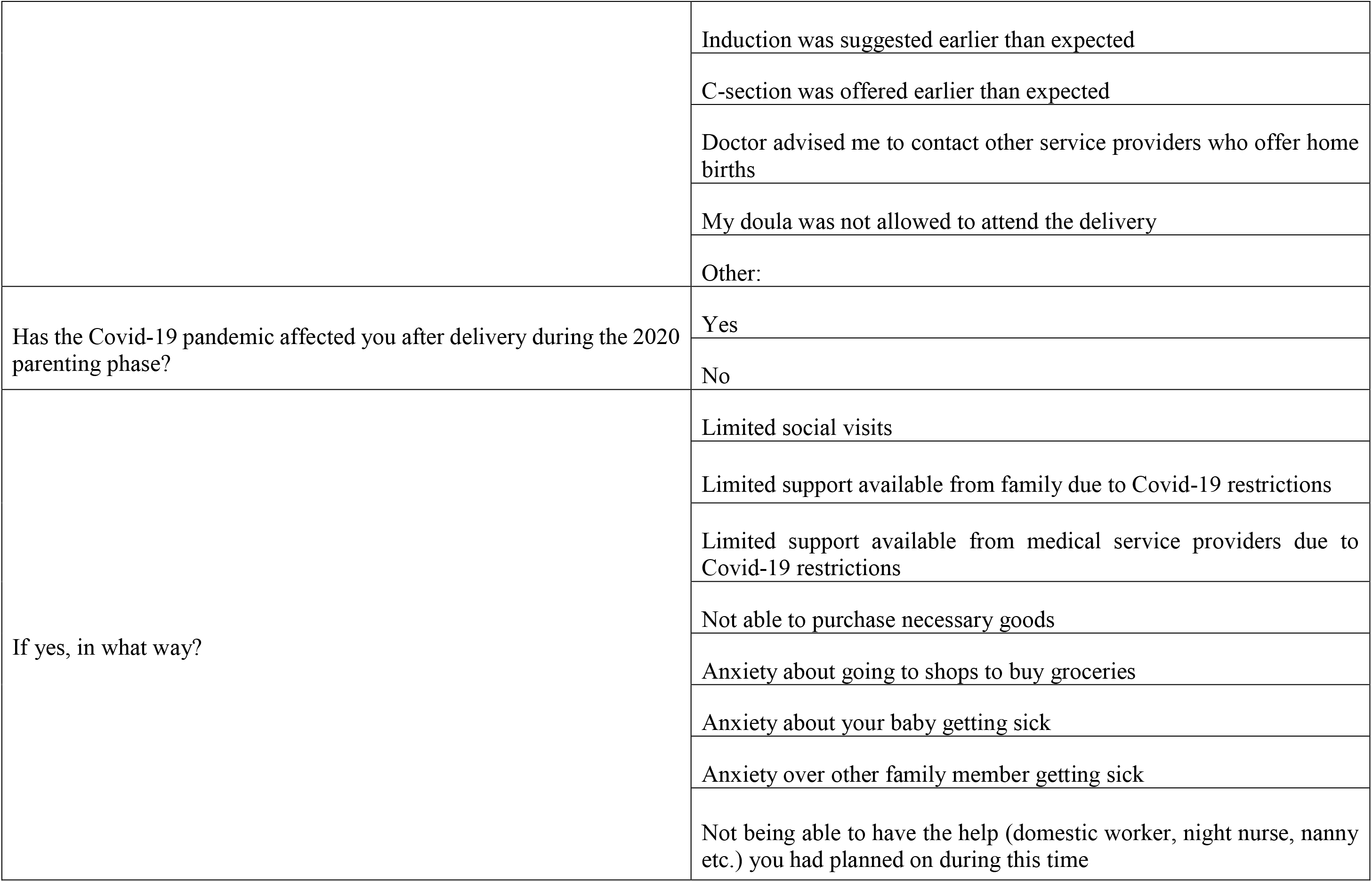
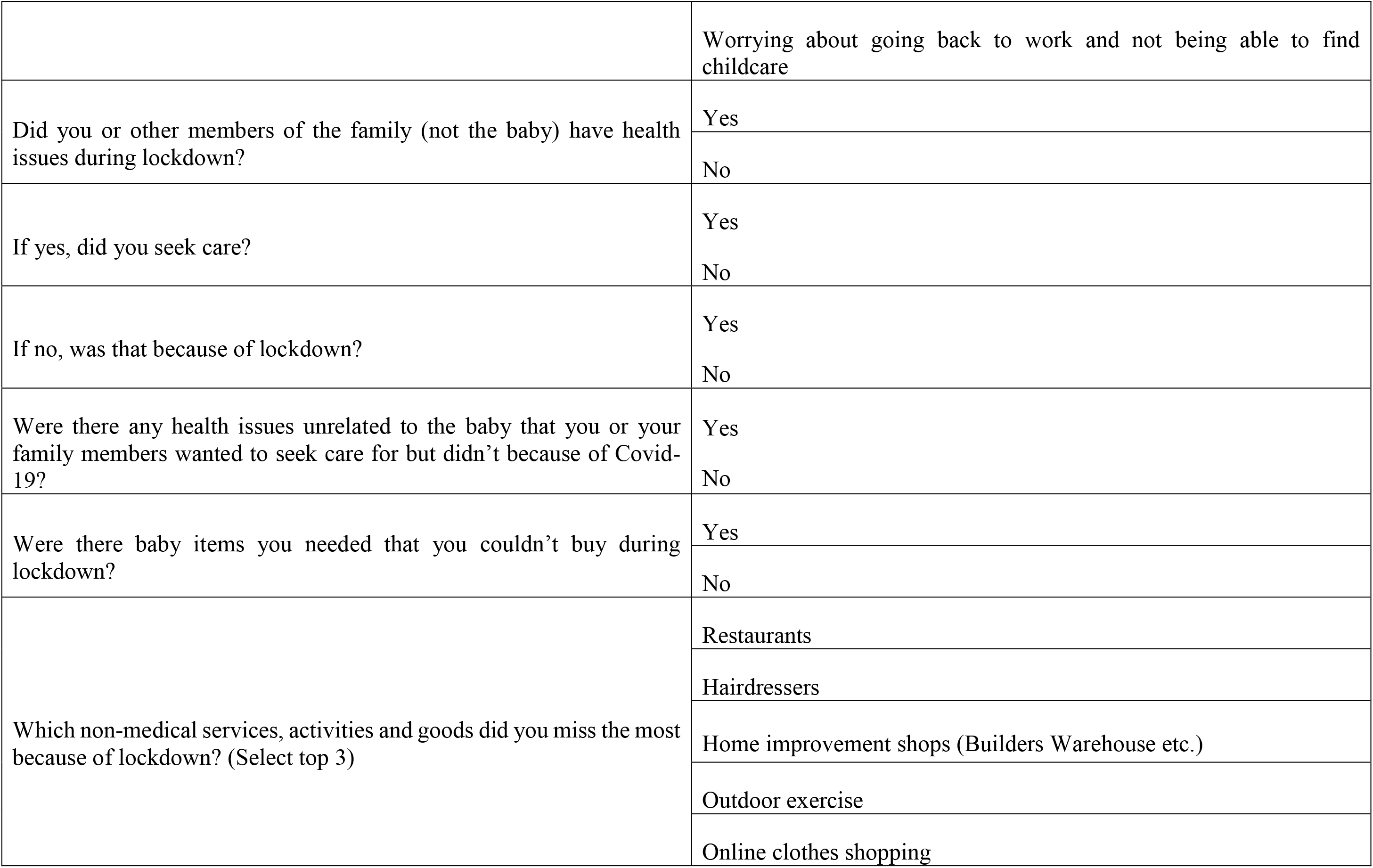
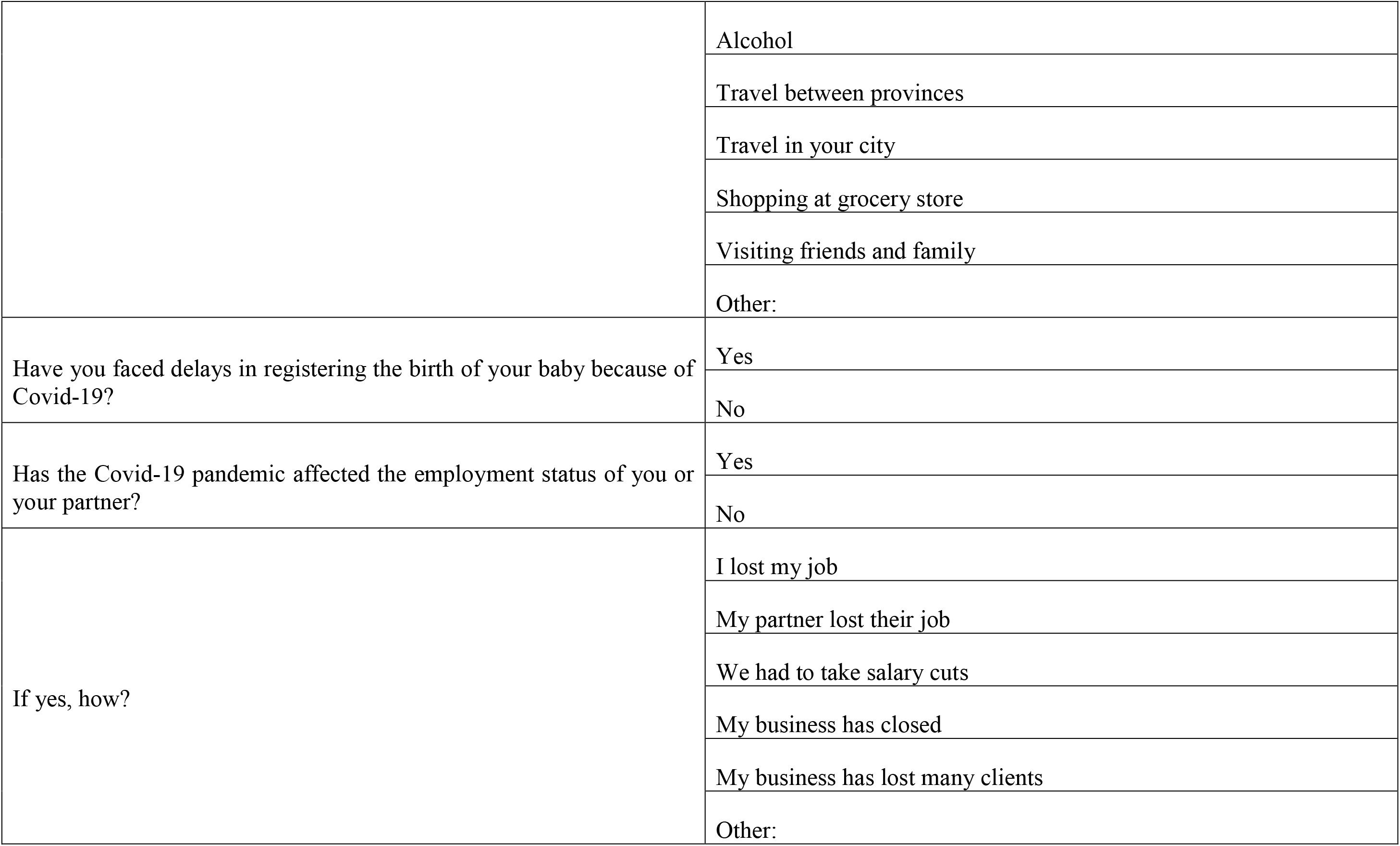
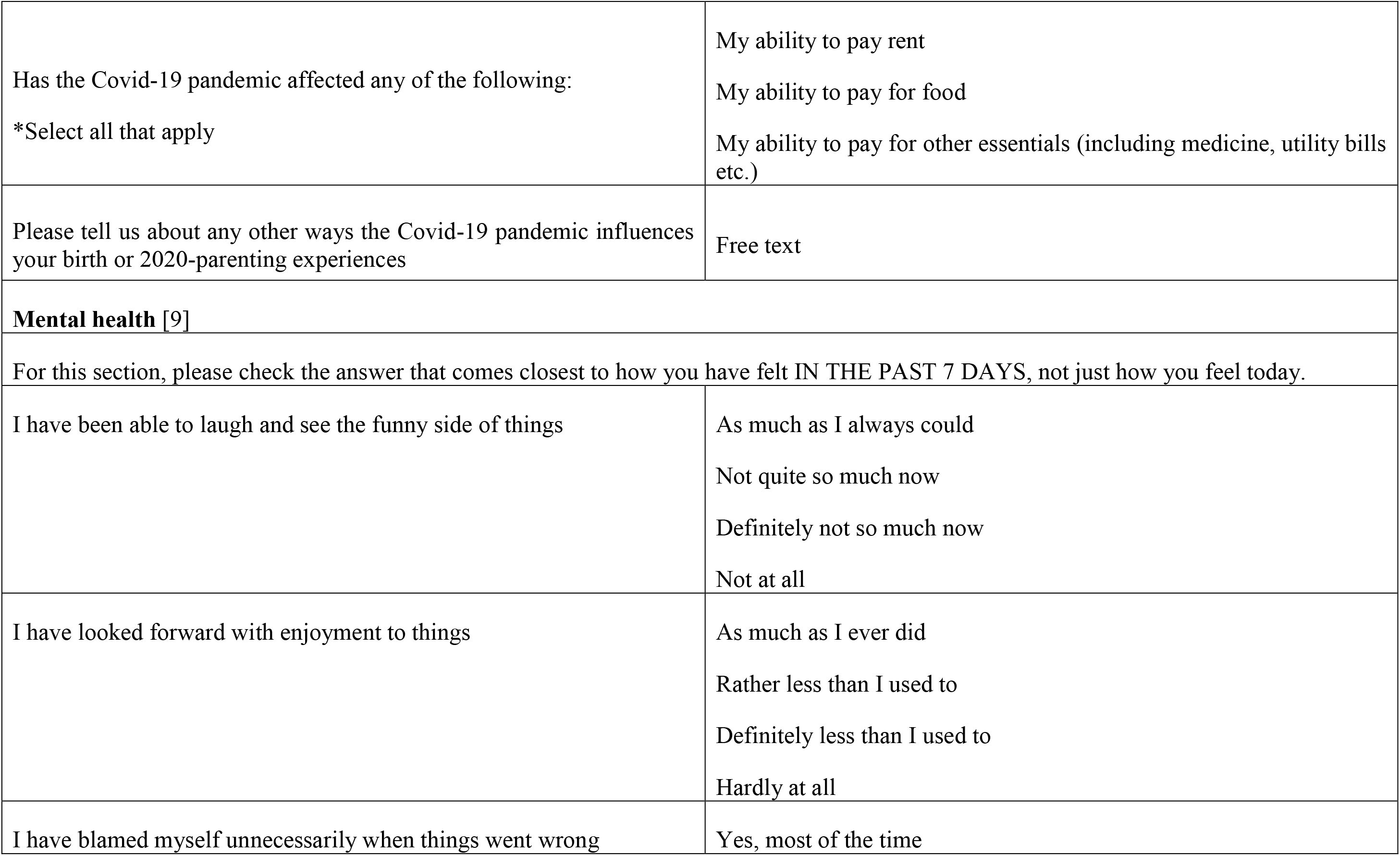
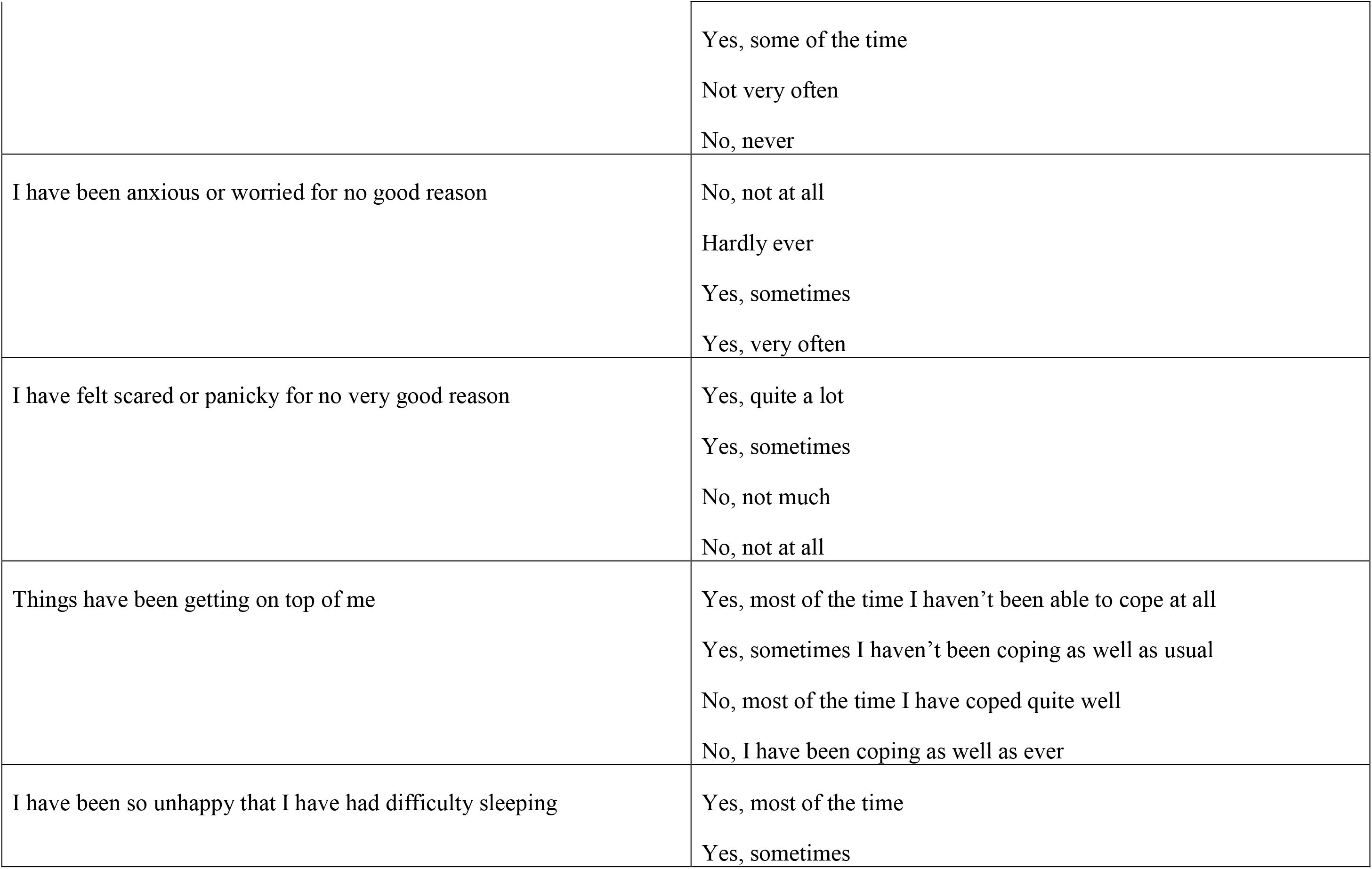
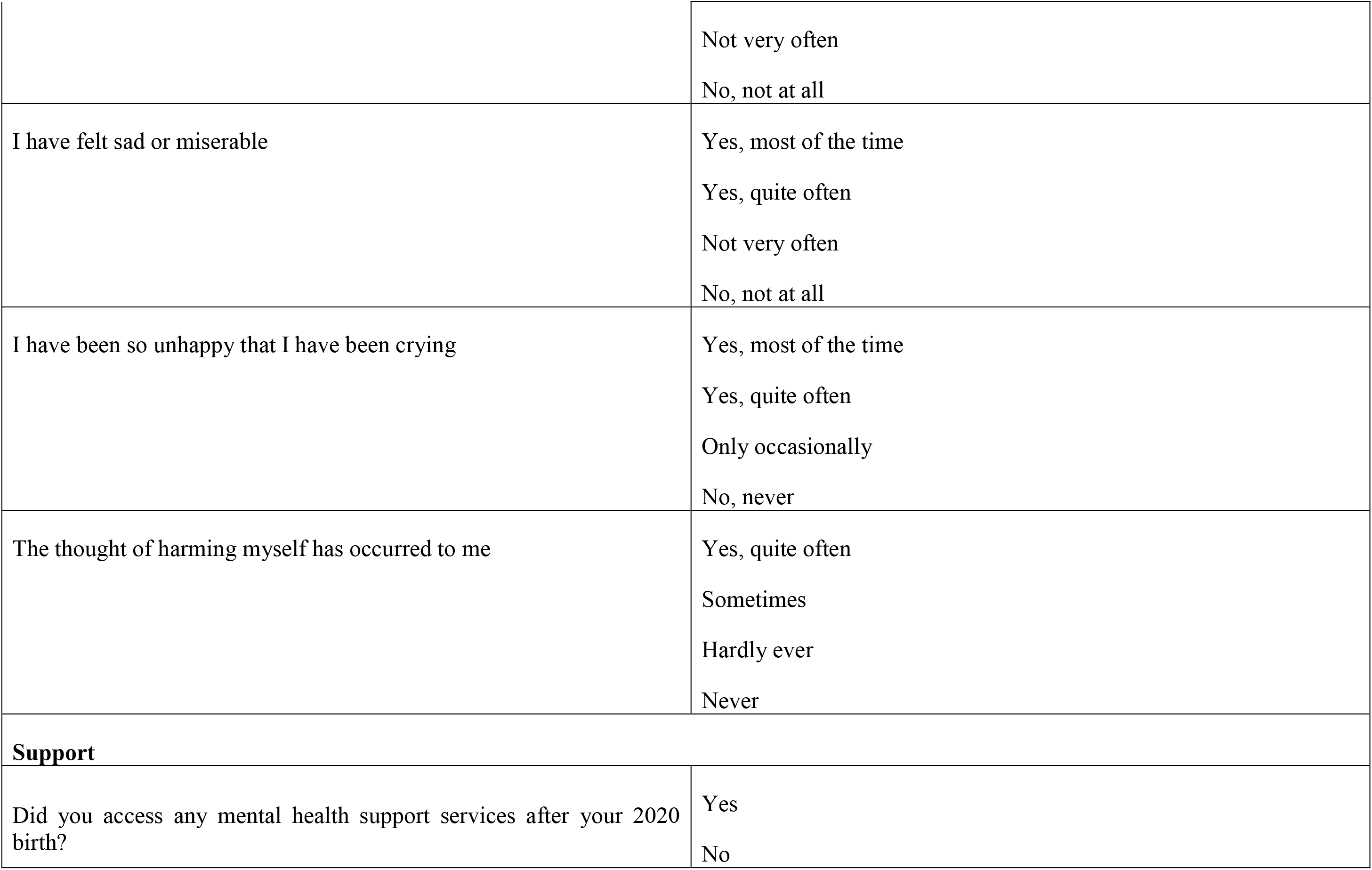
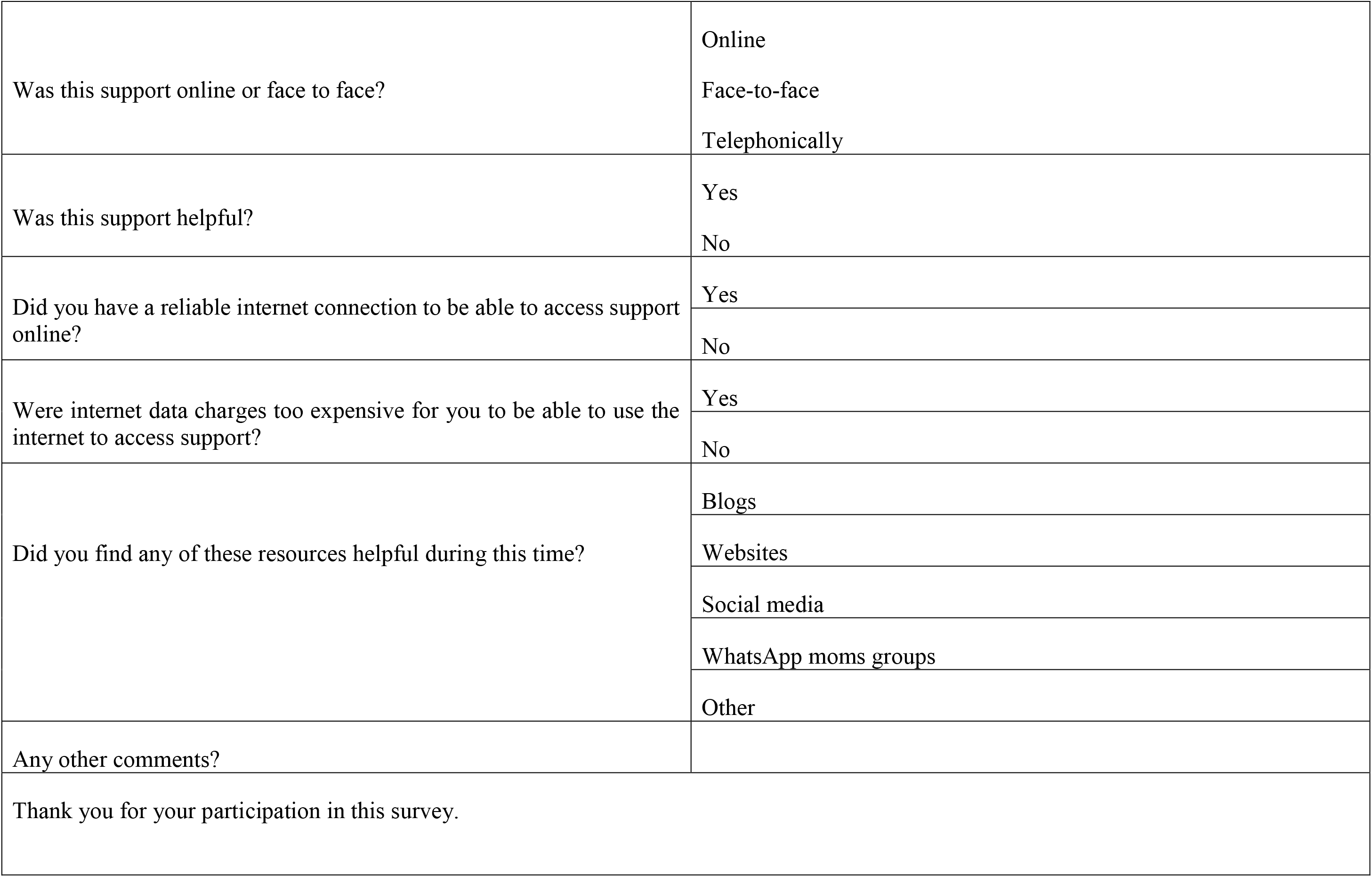
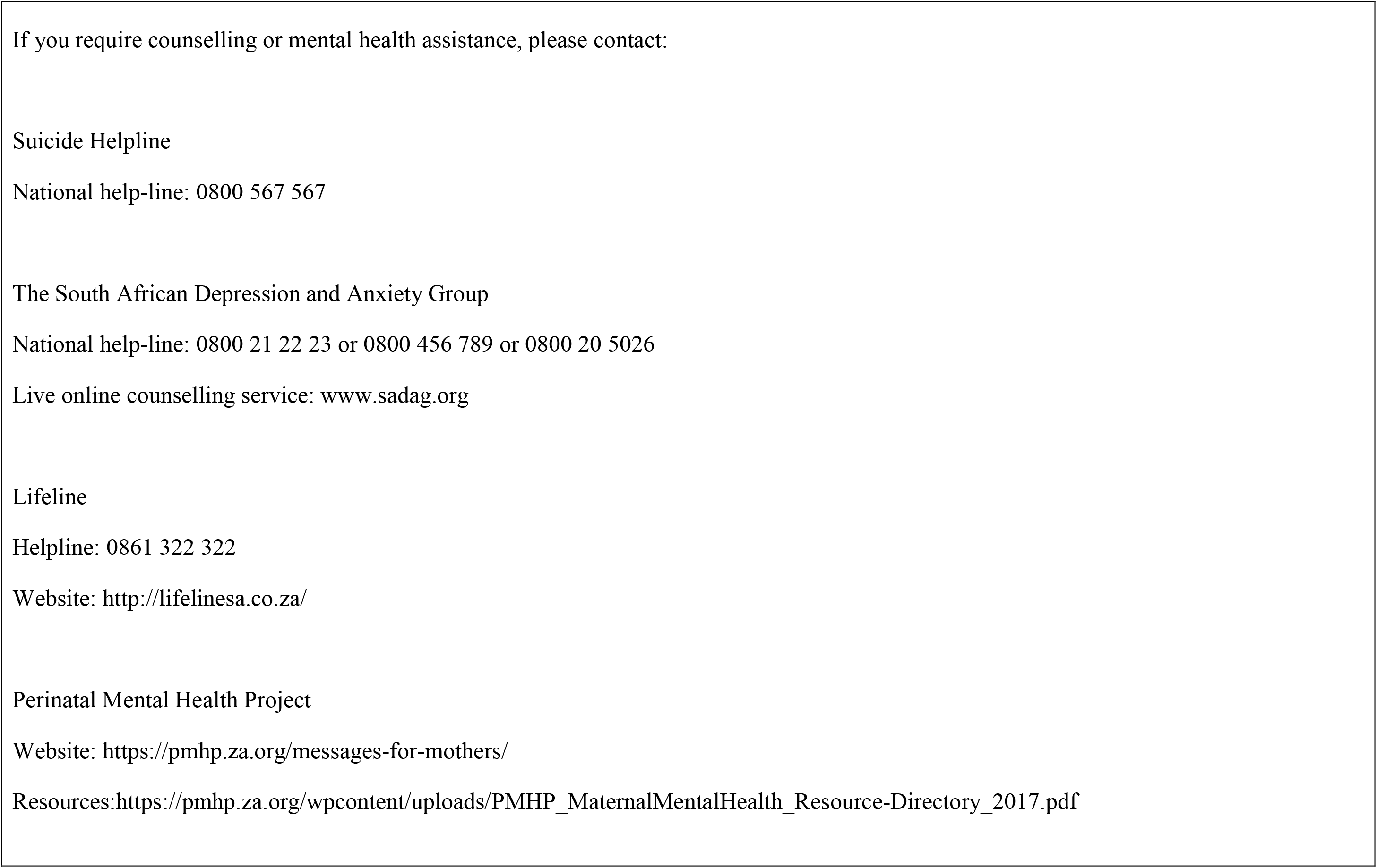
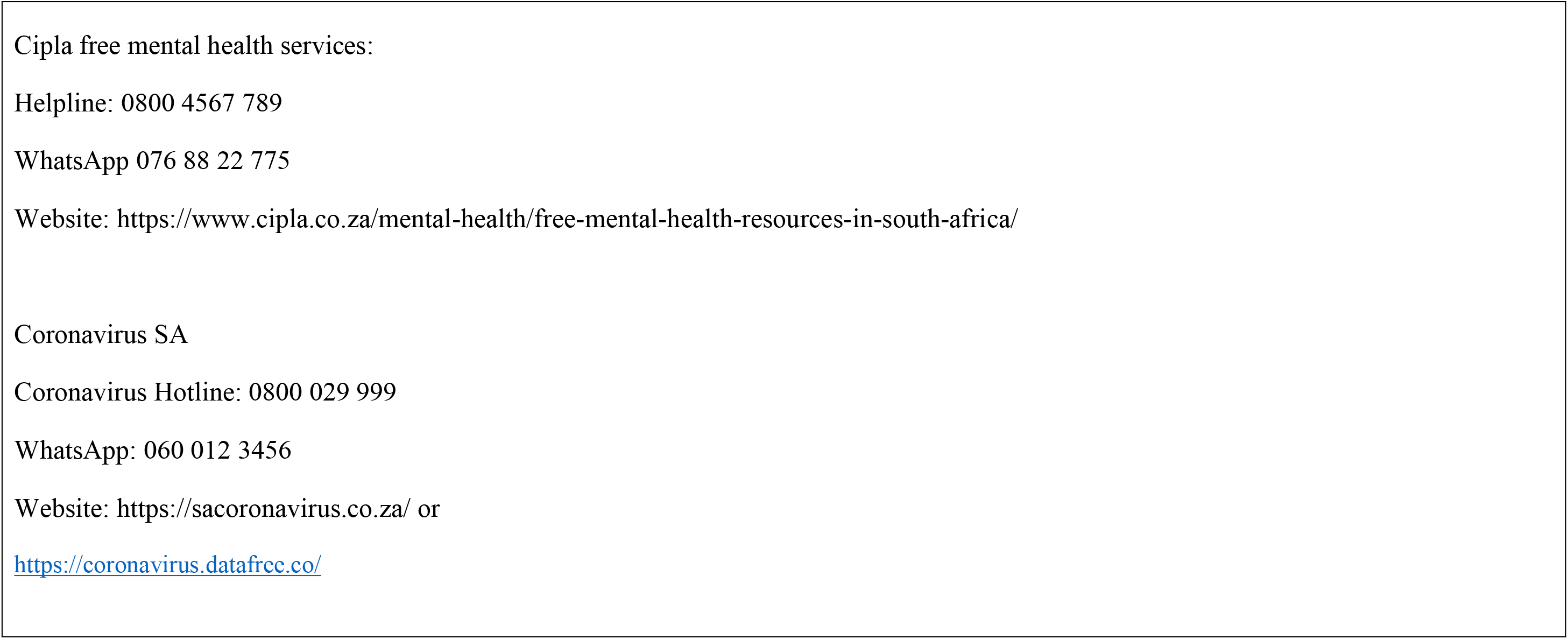
Study Questionnaire

**Supplementary material, Table S2:** Comparison of respondents’ experiences of birth and the new parenting phase between those with babies born prior to lock down and those born during lockdown.

|  |  |  |  |  |  |  |  |  |
| --- | --- | --- | --- | --- | --- | --- | --- | --- |
|  |  | Total |  | Pre-lockdown |  | Lockdown |  | Ch sq |
| Variable | Categories | N=496 | % | N=78 | % | N=413 | % |  |
| BIRTH |  |  |  |  |  |  |  |  |
| Covid-19 affected birth experience | Yes | 341 | 68.75% | 16 | 20.52% | 325 | 77.75% | <0.001 |
|  | No | 155 | 31.25% | 62 | 79.49% | 93 | 22.25% |  |
| How did the lockdown affect your birth experience | My partner could not visit us after the birth | 152 | 30.60% | 1 | 6.25% | 151 | 46.46% | 0.002 |
|  | My birth plan changed due to the pandemic | 60 | 12.10% | 1 | 6.25% | 59 | 18.15% | 0.22 |
|  | My partner was not allowed to be at the birth | 53 | 10.70% | 1 | 6.25% | 52 | 16.00% | 0.29 |
|  | My doula was not allowed to attend the delivery | 52 | 10.50% | 1 | 6.25% | 51 | 15.69% | 0.31 |
|  | C-section was offered earlier than expected | 29 | 5.80% | 1 | 6.25% | 28 | 8.62% | 0.74 |
|  | Induction was suggested earlier than expected | 24 | 4.80% | 1 | 6.25% | 23 | 7.08% | 0.9 |
|  | Delivery method was changed by doctor | 19 | 3.80% | 0 | 0.00% | 19 | 5.57% | 0.32 |
|  | My baby was taken away from me after birth (as I was Covid-19 positive) | 4 | 0.80% | 0 | 0.00% | 4 | 1.23% | 0.66 |
| Birth plan prior to birth (binary) | Vaginal | 334 | 67.34% | 49 | 62.82% | 285 | 68.18% | 0.35 |
|  | C-section | 162 | 32.66% | 29 | 37.18% | 133 | 31.82% |  |
| Preferred method of birth occurred | Yes | 304 | 61.29% | 51 | 65.38% | 253 | 60.53% | 0.42 |
|  | No | 192 | 38.71% | 27 | 34.62% | 165 | 39.47% |  |
| Actual birth method (binary) | Vaginal | 185 | 37.30% | 26 | 33.33% | 159 | 38.04% | 0.43 |
|  | C-section | 311 | 62.70% | 52 | 66.67% | 259 | 61.96% |  |
| Skin to skin after birth | Yes | 380 | 76.61% | 67 | 85.90% | 313 | 74.88% | 0.04 |
|  | No | 116 | 23.39% | 11 | 14.10% | 105 | 25.12% |  |
| Predominant birth emotion | Negative | 97 | 19.56% | 3 | 3.85% | 94 | 22.49% | <0.001 |
|  | Positive | 399 | 80.44% | 75 | 96.15% | 324 | 77.51% |  |
| Feeding plan before birth | Breastfeed | 438 | 88.31% | 65 | 83.33% | 373 | 89.23% | 0.33 |
|  | Formula feed | 22 | 4.44% | 6 | 7.69% | 16 | 3.83% |  |
|  | Mixed (formula and breastfed) | 34 | 6.85% | 7 | 8.97% | 27 | 6.46% |  |
|  | Did not have a preference | 2 | 0.40% | 0 | 0.00% | 2 | 0.48% |  |
| Saw lactation consultant in hospital | Yes | 83 | 16.73% | 30 | 38.46% | 53 | 12.68% | <0.001 |
|  | No | 413 | 83.27% | 48 | 61.54% | 365 | 87.32% |  |
| Gestational age of baby at birth | Premature (0-36 weeks) | 57 | 11.5% | 9 | 11.50% | 48 | 11.50% | 0.91 |
|  | Normal (37-42 weeks) | 436 | 87.9% | 69 | 88.50% | 367 | 87.80% |  |
|  | Prolonged (43 or more weeks) | 1 | 0.2% | 0 | 0.00% | 1 | 0.20% |  |
|  | Missing | 2 | 0.4% | 0 | 0.00% | 2 | 0.50% |  |
| Someone I wanted to be at the birth was not able to be there because of Covid-19 | Yes | 204 | 41.13% | 6 | 7.69% | 198 | 47.37% | <0.001 |
|  | No | 292 | 58.87% | 72 | 92.31% | 220 | 52.63% |  |
| NEW PARENTING |  |  |  |  |  |  |  |  |
| Covid-19 affected me after delivery | Yes | 412 | 83.06% | 67 | 85.90% | 345 | 82.54% | 0.47 |
|  | No | 84 | 16.94% | 11 | 14.10% | 73 | 17.46% |  |
| How Covid-19 affected me after delivery | Limited social visits | 339 | 68.30% | 57 | 85.07% | 282 | 81.74% | 0.51 |
|  | Limited support available from family due to Covid-19 restrictions | 321 | 64.70% | 54 | 80.60% | 267 | 77.39% | 0.56 |
|  | Anxiety about your baby getting sick | 305 | 61.50% | 48 | 71.64% | 257 | 74.49% | 0.63 |
|  | Anxiety about going to shops to buy groceries | 304 | 61.30% | 49 | 73.13% | 255 | 73.91% | 0.89 |
|  | Anxiety over other family member getting sick | 267 | 53.80% | 43 | 64.18% | 224 | 64.93% | 0.91 |
|  | Not being able to have the help (domestic worker, night nurse, nanny etc.) you had planned on during this time | 232 | 46.80% | 49 | 73.13% | 183 | 53.04% | 0.002 |
|  | Worrying about going back to work and not being able to find childcare | 173 | 34.90% | 38 | 56.72% | 135 | 39.13% | 0.008 |
|  | Not able to purchase necessary goods | 126 | 25.40% | 23 | 34.33% | 103 | 29.86% | 0.47 |
|  | Limited support available from medical service providers due to Covid-19 restrictions | 109 | 22.00% | 24 | 35.82% | 85 | 24.64% | 0.06 |
| Predominant new parenting emotions | Negative | 151 | 30.44% | 25 | 32.05% | 126 | 30.14% | 0.74 |
|  | Positive | 345 | 69.56% | 53 | 67.95% | 292 | 69.86% |  |
| Struggled with breastfeeding | Yes | 275 | 55.44% | 40 | 51.28% | 235 | 56.22% | 0.11 |
|  | No | 203 | 40.93% | 32 | 41.03% | 171 | 50.91% |  |
|  | Never breastfed | 18 | 3.63% | 6 | 7.69% | 12 | 2.87% |  |
| Reason for struggling with breastfeeding | My nipples were painful and cracked | 122 | 24.60% | 20 | 25.64% | 102 | 24.40% | 0.55 |
|  | I could not get my baby to latch | 113 | 22.80% | 13 | 16.67% | 100 | 23.92% | 0.37 |
|  | I feel my milk supply is too low | 99 | 20.00% | 13 | 16.67% | 86 | 20.57% | 0.64 |
|  | I did not know if my baby was getting enough milk | 84 | 16.90% | 8 | 10.26% | 76 | 18.18% | 0.23 |
|  | My breasts were engorged | 60 | 12.10% | 7 | 8.97% | 53 | 12.68% | 0.57 |
|  | I have flat or inverted nipples | 43 | 8.70% | 2 | 2.56% | 41 | 9.81% | 0.11 |
|  | Clogged milk ducts | 38 | 7.70% | 10 | 12.82% | 28 | 6.70% | 0.07 |
|  | My baby has an anatomical mouth problem which makes breastfeeding (such as a tongue tie or cleft) | 31 | 6.30% | 5 | 6.41% | 26 | 6.22% | 0.69 |
|  | I got mastitis | 24 | 4.80% | 5 | 6.41% | 19 | 4.55% | 0.49 |
|  | My baby got nipple confusion | 16 | 3.20% | 1 | 1.28% | 15 | 3.59% | 0.47 |
| Conflicting breastfeeding advice | Yes | 209 | 42.14% | 33 | 42.31% | 176 | 42.11% | 0.97 |
|  | No | 287 | 57.86% | 45 | 57.69% | 242 | 57.89% |  |
| **MENTAL HEALTH** |  |  |  |  |  |  |  |  |
| Edinburgh scale (*More sensitive) | Likely no depression (0-9) | 182 | 36.69% | 28 | 35.90% | 154 | 36.84% | 0.87 |
|  | Probable minor depression (10 or more) | 314 | 63.31% | 50 | 64.10% | 264 | 63.16% |  |
| Edinburgh scale | Likely no major depression (0-12) | 282 | 56.85% | 49 | 62.82% | 233 | 55.74% | 0.25 |
|  | Probable major depression (13 or more) | 214 | 43.15% | 29 | 37.18% | 185 | 44.26% |  |

## References

1. Wrottesley S V., Lamper C, Pisa PT. Review of the importance of nutrition during the first 1000 days: Maternal nutritional status and its associations with fetal growth and birth, neonatal and infant outcomes among African women. J Dev Orig Health Dis. 2015;7(2):144–62. DOI: 10.1017/S2040174415001439

2. Garthus-Niegel S, von Soest T, Vollrath M, Eberhard-Gran M. The impact of subjective birth experiences on post-traumatic stress symptoms: a longitudinal study. Arch Womens Ment Heal. 2013;16:1–10. DOI: http://dx.doi.org.ezproxy.uct.ac.za/10.1007/s00737-012-0301-3

3. Garthus-Niegel S, Knoph C, von Soest T, Nielsen CS, Eberhard-Gran M. The Role of Labor Pain and Overall Birth Experience in the Development of Posttraumatic Stress Symptoms: A Longitudinal Cohort Study. Birth. 2014;41(1):108–15. DOI: 10.1111/birt.12093

4. Latva R, Korja R, Salmelin RK, Lehtonen L, Tamminen T. How is maternal recollection of the birth experience related to the behavioral and emotional outcome of preterm infants? Early Hum Dev. 2008;84(9):587–94. DOI: 10.1016/j.earlhumdev.2008.02.002

5. Gutierrez-Galve L, Stein A, Hanington L, Heron J, Ramchandani P. Paternal depression in the postnatal period and child development: Mediators and moderators. Pediatrics. 2015;135(2):e339–47. DOI: 10.1542/peds.2014-2411

6. Fiorillo A, Gorwood P. The consequences of the COVID-19 pandemic on mental health and implications for clinical practice. Eur Psychiatry. 2020;63(1):e32. DOI: 10.1192/j.eurpsy.2020.35

7. Venkatesh A, Edirappuli S. Social distancing in covid-19: What are the mental health implications? BMJ. 2020;369(April):2020. DOI: 10.1136/bmj.m1379

8. Vazquez-Vazquez A, Dib S, Rougeaux E, Wells J, Fewtrell M. The impact of the Covid-19 lockdown on the experiences and feeding practices of new mothers in the UK: Preliminary data from the COVID-19 New Mum Study. medRxiv. 2020; DOI: doi.org/10.1101/2020.06.17.20133868

9. Cox J, Holden J, Sagovsky R. Detection of postnatal depression: Development of the 10-item Edinburgh Postnatal Depression Scale. Br J Psychiatry 150782-786. 1987;150:782–6.

10. Matthey S, Henshaw C, Elliott S, Barnett B. Variability in use of cut-off scores and formats on the Edinburgh Postnatal Depression Scale - Implications for clinical and research practice. Arch Womens Ment Health. 2006;9(6):309–15. DOI: 10.1007/s00737-006-0152-x

11. Chen Q, Nian H, Zhu Y, Talbot H, Griffin M, Harrell F. Too many covariates and too few cases? – A comparative study. Stat Med. 2017;35(25):4546–58. DOI: 10.1002/sim.7021.Too

12. WHO. Preterm birth: Key facts [Internet]. World Health Organization. 2018 [cited 2021 May 8].

13. Stampini V, Monzani A, Amadori R. A Survey Among Italian Pregnant Women and New-mothers During the COVID-19 Pandemic Lockdown. :1–16.

14. Cameron EE, Joyce KM, Rollins K, Roos LE. Paternal Depression & Anxiety During the COVID-19 Pandemic. Preprint. 2020; DOI: 10.31234/osf.io/drs9u

15. Matthews S. More Midwife-based Interventions Could Save Millions of Lives. New Security Beat. 2020 Dec 9;

16. Gibson L, Hanson VL. “Digital motherhood”: How does technology support new mothers? Conf Hum Factors Comput Syst - Proc. 2013;313–22. DOI: 10.1145/2470654.2470700

17. Rabkin F. Products for the care of babies and toddlers now essential items. Mail and Guardian. 2020 Apr 6;

18. Alvar J, Vélez ID, Bern C, Herrero M, Desjeux P, Cano J, Jannin J, de Boer M. Leishmaniasis worldwide and global estimates of its incidence. PLoS One. 2012;7(5). DOI: 10.1371/journal.pone.0035671

19. R.P. N, J. M. Rising rates of Caesarean sections: An audit of Caesarean sections in a specialist private practice. South African Fam Pract. 2009;51(3):254–8.

20. Solanki GC, Cornell JE, Daviaud E, Fawcus S. Caesarean section rates in South Africa: A case study of the health systems challenges for the proposed National Health Insurance. South African Med J. 2020;110(8):747–50. DOI: 10.7196/SAMJ.2020.v110i8.14699

21. Council for Medical Schemes. Epidemiology and trends of caesarean section births in the medical schemes’. 2020.

22. World Health Organisation. WHO statement on caesarean section rates. World Health Organisation. 2015. DOI: 10.1111/1471-0528.13526

23. Benton M, Salter A, Tape N, Wilkinson C, Turnbull D. Women’s psychosocial outcomes following an emergency caesarean section: A systematic literature review. BMC Pregnancy Childbirth. 2019;19(1). DOI: 10.1186/s12884-019-2687-7

24. Chen H, Tan D. Cesarean section or natural childbirth? Cesarean birth may damage your health. Front Psychol. 2019;10(FEB):1–7. DOI: 10.3389/fpsyg.2019.00351

25. Hobbs AJ, Mannion CA, McDonald SW, Brockway M, Tough SC. The impact of caesarean section on breastfeeding initiation, duration and difficulties in the first four months postpartum. BMC Pregnancy Childbirth. 2016;16(1):1–9. DOI: 10.1186/s12884-016-0876-1

26. Stinson LF, Payne MS, Keelan JA. A critical review of the bacterial baptism hypothesis and the impact of cesarean delivery on the infant microbiome. Front Med. 2018;5(MAY). DOI: 10.3389/fmed.2018.00135

27. Etikan I, Musa S, Alkassim R. Comparison of convenience sampling and purposive sampling. Am J Theor Appl Stat. 2016;5(1):1–4. DOI: 10.11648/j.ajtas.20160501.11

